# IMPACT OF BACKGROUND IGBO HIGHLIFE MUSIC ON COGNITIVE PERFORMANCE AMONG CLINICAL MEDICAL STUDENTS: A COMPARATIVE STUDY OF MEMORY RECALL AND PROBLEM-SOLVING EFFICIENCY

**DOI:** 10.64898/2026.02.22.26346677

**Authors:** Chidiebere Jude Anaenye, Azuoma Lasbrey Asomugha

## Abstract

**Background:** The cognitive demands of medical education require optimal learning environments. While the influence of background music on cognition has been widely studied, existing research exhibits a significant Eurocentric bias, predominantly focusing on Western classical music like the “Mozart Effect.” This leaves a critical gap in understanding the impact of culturally salient, non-Western musical traditions on learning within their native contexts.

**Methods:** A single-blind, randomized controlled trial was conducted with 147 clinical (4th, 5th and 6th) year medical students stratified by ethnicity at the Faculty of Medicine, Nnamdi Azikiwe University, Nnewi Campus, Anambra State, Nigeria between March and September 2025. Participants were randomly assigned to one of three background music conditions: Igbo Highlife (instrumental), Western classical (Mozart’s Sonata K.448), or silence (control with pink noise masking). Cognitive performance was assessed through a short-term memory recall test of 20 medical terms and a timed clinical problem-solving task comprising 20 multiple-choice questions. Baseline mood was controlled for using the Positive and Negative Affect Schedule (PANAS). Data were analyzed using ANOVA and post-hoc Tukey HSD tests.

**Results:** Music condition had a highly significant effect on student performance (p < 0.001). The Igbo Highlife group demonstrated superior outcomes, achieving the highest scores in memory recall (mean = 16.7) and problem-solving accuracy (mean = 15.7), alongside the fastest completion time (23.4 seconds/question), significantly outperforming both the classical and silence groups. A significant correlation was found between cultural familiarity with Highlife and enhanced cognitive performance (accuracy: ρ = 0.268, p = 0.001).

**Conclusions:** Incorporating music that holds cultural significance and familiarity to learners, specifically Igbo Highlife, is a highly effective auditory stimulus for enhancing learning efficiency in medical education. Students and educational institutions should consider integrating culturally familiar instrumental music into study environments to optimize cognitive performance and learning outcomes.

**Competing Interest Statement:** The authors have declared no competing interest.

## INTRODUCTION

Cognitive performance in high-stakes academic environments, such as medical education, is not merely a product of innate ability but is significantly modulated by the learning environment, including its auditory dimension. Within this context, the strategic use of background music has emerged as a potential cognitive ergonomic tool. The theoretical premise is grounded in the understanding that auditory stimuli can influence neurocognitive states, thereby affecting processes critical to learning, such as attention, memory encoding, and problem-solving [1]. For medical students, who operate under chronic academic pressure and must master vast, complex bodies of declarative and procedural knowledge, identifying environmental strategies to optimize cognitive efficiency is of paramount practical importance.

The scientific discourse on music and cognition was profoundly shaped by the discovery of the “Mozart Effect,” which reported temporary enhancements in spatial-temporal reasoning following exposure to a Mozart sonata [2]. This finding catalyzed a prolific field of inquiry, yet one that has exhibited a persistent and consequential bias. The overwhelming majority of subsequent research has focused on Western populations and a narrow canon of Western art music, most notably compositions by Mozart and other classical-era figures [3]. This corpus of work has elucidated key psychological mechanisms through which music may operate, primarily articulated through two complementary theoretical lenses. The Arousal-Mood Hypothesis posits that music acts as a modulator of affective and physiological states, potentially reducing stress-induced cognitive interference and elevating mood to an optimal level for task engagement [4]. Concurrently, Cognitive Load Theory provides a framework for understanding how auditory stimuli interact with finite cognitive resources, suggesting that instrumental music may reduce extraneous load by masking disruptive ambient noise, thereby freeing capacity for germane, learning-relevant processing [5].

However, this dominant paradigm harbors a critical, often unexamined assumption: that the cognitive effects of music are universal, transcending cultural context. This assumption is directly challenged by the Theory of Enculturation, which holds that musical perception is not innate but is sculpted by prolonged exposure to the specific rhythmic, melodic, and harmonic syntax of one’s native soundscape [6]. From this perspective, a familiar musical genre is processed with greater neural efficiency, requiring less cognitive effort for decoding and integration than an unfamiliar one. Consequently, the purported benefits of Western classical music for Western listeners may stem as much from this cultural fluency as from any inherent properties of the music itself.

This gap is especially pronounced and practically relevant within African educational landscapes. In Nigeria, a nation with profound cultural and musical diversity, students’ auditory worlds are richly populated by indigenous genres whose structures and affective connotations differ radically from Western classical forms. Igbo Highlife is a prime exemplar. Originating in southeastern Nigeria in the mid-20th century, Highlife synthesizes traditional Igbo melodic and rhythmic motifs with imported instruments, creating an upbeat, polyrhythmic, and deeply resonant popular genre [7]. For the Igbo people and many other Nigerians, Highlife is more than entertainment; it is an integral component of cultural identity and social life, intrinsically linked to communal gatherings and positive emotional experiences. This profound cultural salience makes it an ideal candidate for testing the enculturation hypothesis in a cognitive setting. Its rhythmic complexity may support neural entrainment and sustained attention, while its deep familiarity for the target population should, theoretically, minimize the extraneous cognitive load associated with processing an unfamiliar auditory stream. Despite its cultural centrality, the potential cognitive impact of Highlife, and of African music more broadly, remains virtually unexplored in controlled experimental literature.

At the time of this study, no randomized controlled trial in Africa had investigated the cognitive effects of culturally familiar music on medical students. Consequently, three specific gaps remained unaddressed: (1) the effect of indigenous African music, such as Igbo Highlife, on cognitive performance in a controlled experiment; (2) a direct comparison of its efficacy for medical memory recall and problem-solving against both Western classical music and silence; and (3) an assessment of whether cultural familiarity moderates these cognitive outcomes. This lack of localized evidence presents a practical problem: students and educators cannot make informed decisions about using music as a cognitive aid in non-Western educational contexts.

This study was therefore designed to address these gaps and provide an empirical foundation for optimizing study environments in Nigerian medical education. The aim of this study was to evaluate the impact of Igbo Highlife background music on cognitive performance in clinical medical students. The specific objectives were: (1) to compare the effect of Igbo Highlife background music on memory recall against Western classical music and silence; (2) to evaluate the influence of Igbo Highlife background music on the speed and accuracy of problem-solving in clinical tasks; and (3) to determine the correlation between the level of cultural familiarity with Igbo Highlife music and cognitive performance outcomes.

We hypothesized that medical students exposed to Igbo Highlife background music would demonstrate significantly better memory recall and problem-solving performance than those exposed to Western classical music or silence, and that there would be a significant positive correlation between cultural familiarity with Igbo Highlife music and cognitive performance outcomes.

## MATERIALS AND METHODS

### Study Design

This study employed a single-blind, randomized controlled trial with a three-parallel group design. Participants were randomly assigned to one of three auditory conditions: Igbo Highlife music, Western classical music, or silence (control). The analyst was blinded to group assignment. Testing included both immediate and delayed (48-hour) components.

### Ethical Approval

Ethical approval for this study was obtained from the Nnamdi Azikiwe University Teaching Hospital Health Research Ethics Committee (Approval Number: NAUTH/CS/66/VOL.18/VER.3/141/2025/48). All participants provided written informed consent prior to enrollment. The study was conducted in accordance with the Declaration of Helsinki.

### Study Setting and Location

The experimental sessions were conducted at the Gilbert Metu Uzodike Auditorium, Nnamdi Azikiwe University Teaching Hospital, Nnewi, Anambra State, Nigeria, between March and September 2025. The venue provided a large, controlled environment with uniform lighting and minimal external noise. Participants were spaced at intervals to prevent auditory interference, and all audio stimuli were delivered via closed-back, noise-isolating headphones (Bose QC45).

### Participants

#### Eligibility Criteria

Participants were eligible for inclusion if they were: (1) enrolled in the clinical years (4th, 5th, or 6th year) of the medical program at Nnamdi Azikiwe University; and (2) had self-reported normal or corrected-to-normal hearing and vision. Exclusion criteria were: (1) self-reported hearing impairment; and (2) a prior diagnosis of a cognitive or learning disability.

#### Sample Size Calculation

An a priori power analysis was conducted using G*Power 3.1 for a one-way analysis of variance. With an alpha level of 0.05, a power of 0.80, and a medium effect size (η² = 0.15), the minimum required sample size was calculated as 96 participants. To accommodate an anticipated attrition rate of 20%, a total of 175 participants were recruited.

#### Final Sample

Of the 175 participants recruited, 28 were excluded due to incomplete attendance or protocol deviation. The final analyzed sample consisted of 147 participants, randomly allocated as follows: Igbo Highlife group (n = 56), Western Classical group (n = 57), and Silence group (n = 34).

#### Randomization and Blinding

Participants were stratified by ethnicity (Igbo vs. non-Igbo) and then randomly assigned to one of the three auditory conditions using a computer-generated random number sequence. Group allocation was concealed from participants until the commencement of the intervention on Day 2. The analyst responsible for data processing and statistical analysis was blinded to group assignment throughout the study.

### Variables

#### Independent Variable

The independent variable was the auditory condition, with three levels: (1) Igbo Highlife music; (2) Western classical music; and (3) silence (control).

#### Dependent Variables

The primary dependent variables were:

1. **Memory Recall Score:** The number of correctly recalled items from a 20-term medical word list (expressed as raw score).
2. **Problem-Solving Accuracy:** The number of correctly answered clinical multiple-choice questions (expressed as raw score).
3. **Problem-Solving Speed:** The mean time (in seconds) taken per question to complete the 20 clinical vignettes.

#### Moderating Variables

The following moderating variables were measured:

1. **Cultural Familiarity:** Operationalized as the mean score from a 5-point Likert scale (1 = Not at all familiar, 5 = Extremely familiar) assessing self-reported recognition and listening frequency of Igbo Highlife music.
2. **Baseline Affect:** Measured using the Positive and Negative Affect Schedule [8] administered prior to the intervention.
3. **Musical Training:** A dichotomous variable (Yes/No) based on self-report of formal musical training for two or more years.

### Materials and Apparatus

#### Music Stimuli

Three auditory conditions were employed:

- **Igbo Highlife:** A 5-minute instrumental excerpt from Chief Stephen Osita Osadebe’s “Ebezina,” selected for its cultural salience and absence of lyrics.
- **Western Classical:** A standardized 5-minute excerpt from Mozart’s Sonata for Two Pianos in D Major, K. 448, widely used in music-cognition research [2].
- **Silence (Control):** No audio stimulus; participants wore noise-isolating headphones without audio input.

All music tracks were normalized to -16 LUFS (Loudness Units Full Scale) using Audacity software to ensure consistent perceived volume across conditions. Stimuli were delivered via Bose QC45 noise-isolating headphones.

### Cognitive Assessment Tasks

- **Memory Recall Task:** A 20-item list of complex medical terms was developed and piloted for equivalence in difficulty. Participants studied the list for 5 minutes while exposed to their assigned auditory condition. Immediate recall was tested after a 2-minute non-verbal distraction task. Delayed recall was tested 48 hours later using the same word list.
- **Problem-Solving Task:** Twenty clinical multiple-choice questions based on realistic patient vignettes were developed from standardized medical education resources. Questions were designed to assess clinical reasoning and were required to be completed within a 15-minute time limit.

### Affective Measure

Baseline mood states were assessed using the Positive and Negative Affect Schedule (PANAS) [8], a 20-item self-report measure with established reliability and validity.

### Procedure

The study procedure unfolded over three consecutive sessions:

#### Day 1 (Baseline Assessment)

Participants provided written informed consent, completed a demographic questionnaire (including items on ethnicity, cultural familiarity with Igbo Highlife, and musical training), and completed the PANAS to establish baseline affect.

#### Day 2 (Intervention and Immediate Testing)

Participants were randomly assigned to one of the three auditory conditions. They listened to their assigned audio track (or silence) for 5 minutes while studying the 20-item medical term list. Following the study period, the audio ceased, and participants completed a 2-minute non-verbal distraction task. Immediately thereafter, they completed the free-recall test for the medical terms.

#### Day 3 (Delayed Testing – 48 Hours Later)

Participants returned to the same venue. Without exposure to any music stimulus, they completed the 20-item clinical problem-solving task under timed conditions (15-minute limit).

#### Debriefing

Following completion of all testing sessions, participants were provided with a full debriefing on the study’s purpose, hypotheses, and the specific auditory conditions to which they were assigned.

### Statistical Analysis

Data were analyzed using SPSS (Version 28) and JASP. Descriptive statistics were calculated for all variables. Normality of distribution was assessed using the Shapiro-Wilk test, and homogeneity of variances was assessed using Levene’s test.

#### Primary Analysis

One-way analyses of variance were conducted to compare the effect of auditory condition on each dependent variable: memory recall, problem-solving accuracy, and problem-solving speed. Where significant main effects were detected, post-hoc pairwise comparisons were performed using Tukey’s Honestly Significant Difference test to control for Type I error.

#### Secondary Analysis

A two-way ANOVA was conducted to examine the interaction between auditory condition and cultural familiarity (dichotomized as high vs. low) on cognitive performance. Spearman’s rank correlation was used to examine relationships between cultural familiarity and cognitive outcomes.

Statistical significance was set at p < 0.05 for all analyses. Effect sizes are reported as partial eta-squared (η²p) for ANOVA.

## RESULTS

### Participant Flow and Baseline Characteristics

A total of 175 medical students were recruited for this study. Following exclusions due to incomplete attendance or protocol deviation (n = 28), the final analyzed sample comprised 147 participants. Figure 1 presents the CONSORT flow diagram illustrating participant progression through the study.

**Figure 1:**
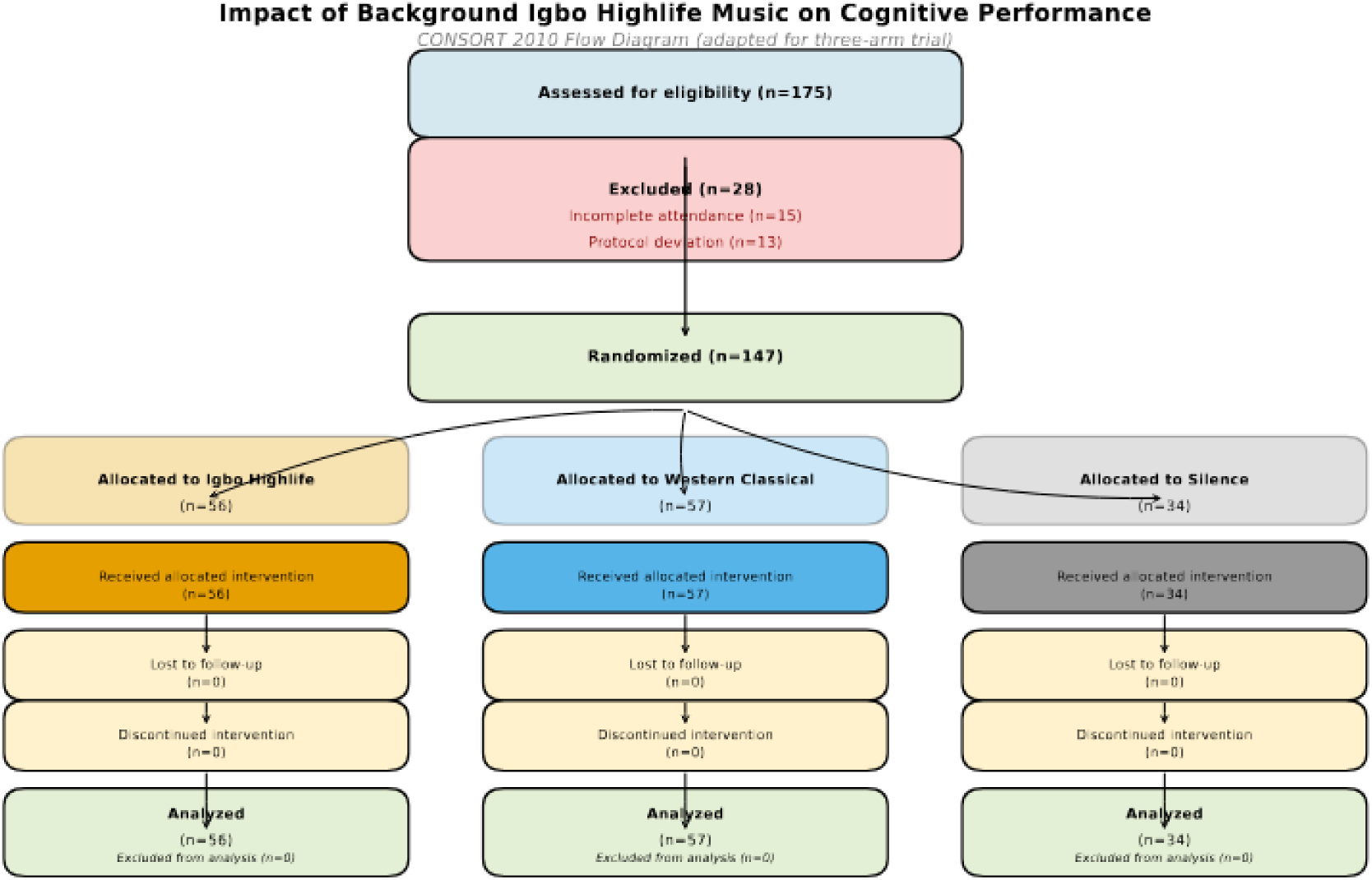
CONSORT Flow Diagram of Participant Progress Through the Study. CONSORT flow diagram showing: Enrollment (175 assessed), Exclusion (28 excluded: 15 incomplete attendance, 13 protocol deviation), Randomization (147 allocated), Allocation (56 to Highlife, 57 to Classical, 34 to Silence), Follow-up, and Analysis]

### Demographic Characteristics

The demographic characteristics of the final sample are presented in Table 1. The majority of participants were female (n = 89, 60.5%), reflecting the demographic profile of the participating medical school. The mean age was 22.01 years (SD = 1.44), and participants reported studying for an average of 2.24 hours per day (SD = 0.81). Self-reported familiarity with Igbo Highlife music demonstrated adequate variation across the sample to permit correlational analyses.

**Table 1:** Demographic and Baseline Characteristics of Participants (N = 147)

| Characteristic | Category | n | % | Mean | SD |
| --- | --- | --- | --- | --- | --- |
| **Gender** | Male | 58 | 39.5 |  |  |
|  | Female | 89 | 60.5 |  |  |
| **Age (years)** |  |  |  | 22.01 | 1.44 |
| **Daily Study Hours** |  |  |  | 2.24 | 0.81 |

### Baseline Mood Assessment

The Positive and Negative Affect Schedule was administered prior to the intervention to assess baseline mood states. A one-way ANOVA confirmed no statistically significant differences between experimental groups in either Positive Affect (F(2,144) = 1.12, p = 0.330) or Negative Affect (F(2,144) = 0.87, p = 0.422) scores at baseline, indicating successful randomization (Table 2). This confirms that any subsequent differences in cognitive performance can be attributed to the experimental manipulation rather than pre-intervention mood variances.

**Table 2:** Baseline PANAS Scores by Experimental Condition (Mean ± SD)

| Condition | n | Positive Affect (PA) Score | Negative Affect (NA) Score |
| --- | --- | --- | --- |
| **Igbo Highlife** | 56 | 20.93 ± 1.54 | 12.79 ± 0.89 |
| **Western Classical** | 57 | 20.61 ± 1.38 | 12.96 ± 0.83 |
| **Silence** | 34 | 20.29 ± 1.62 | 13.12 ± 1.01 |
| **Total** | **147** | **20.65 ± 1.52** | **12.93 ± 0.90** |
| ***p-value*** |  | *0.330* | *0.422* |

### Effect of Auditory Condition on Memory Recall Descriptive Statistics

Table 3 presents the descriptive statistics for memory recall scores across the three auditory conditions. Participants in the Igbo Highlife condition achieved the highest mean recall score (16.7 ± 0.904), followed by the Western Classical condition (13.7 ± 0.630), while the Silence condition produced the lowest mean score (10.2 ± 0.878). These patterns are visualized in Figure 2.

**Figure 2:**
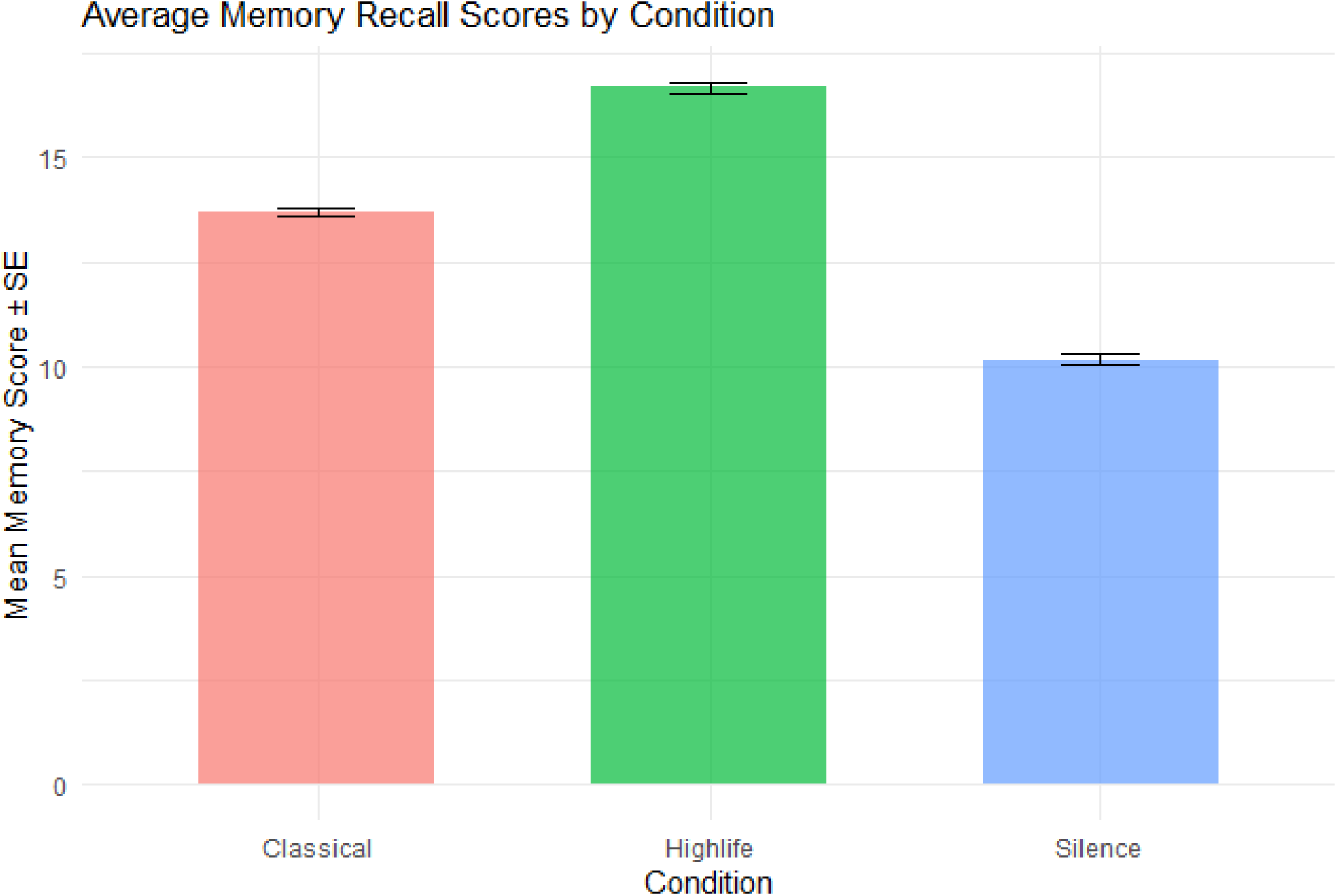
Mean Memory Recall Scores by Auditory Condition Inferential Statistics. A one-way ANOVA revealed a significant main effect of auditory condition on memory recall scores (F(2,144) = 717.8, p < 0.001, η²p = 0.91), indicating that the type of background music substantially influenced memory performance (Table 4).

**Table 3:** Memory Recall Scores by Auditory Condition.

| Condition | n | Mean | SD | 95% CI |
| --- | --- | --- | --- | --- |
| **Igbo Highlife** | 55 | 16.7 | 0.904 | [16.46, 16.94] |
| **Western Classical** | 56 | 13.7 | 0.630 | [13.53, 13.87] |
| **Silence** | 36 | 10.2 | 0.878 | [9.90, 10.50] |

**Table 4:** One-Way ANOVA for Memory Recall Scores by Condition.

| Source | df | Sum of Squares | Mean Square | F | p | $\eta^2p$ |
| --- | --- | --- | --- | --- | --- | --- |
| **Condition** | 2 | 926.6 | 463.3 | 717.8 | <0.001 | 0.91 |
| **Residuals** | 144 | 92.9 | 0.6 |  |  |  |

Post-hoc comparisons using Tukey’s HSD test (Table 5) demonstrated that all pairwise differences were statistically significant (p < 0.001). The Igbo Highlife condition significantly outperformed both the Western Classical condition (mean difference = 2.98, 95% CI [2.62, 3.34]) and the Silence condition (mean difference = 6.51, 95% CI [6.10, 6.91]). The Western Classical condition also significantly outperformed Silence (mean difference = 3.53, 95% CI [3.12, 3.94]).

**Table 5:** Tukey’s HSD Post-Hoc Comparisons for Memory Recall.

| Comparison | Mean Difference | 95% CI Lower | 95% CI Upper | p |
| --- | --- | --- | --- | --- |
| **Highlife – Classical** | 2.98 | 2.62 | 3.34 | <0.001 |
| **Silence – Classical** | -3.53 | -3.94 | -3.12 | <0.001 |
| **Silence – Highlife** | -6.51 | -6.91 | -6.10 | <0.001 |

### Effect of Auditory Condition on Problem-Solving Performance

#### Descriptive Statistics

Table 6 presents descriptive statistics for problem-solving accuracy (MCQ scores) and speed (time per question) across the three auditory conditions. Participants in the Igbo Highlife condition achieved the highest accuracy (15.7 ± 0.855) and the fastest completion time (23.4 ± 1.54 seconds/question). The Western Classical condition yielded intermediate performance (accuracy: 12.6 ± 0.587; time: 27.7 ± 1.03 seconds), while the Silence condition produced the lowest accuracy (7.74 ± 0.710) and slowest completion time (39.6 ± 1.63 seconds). These patterns are visualized in Figures 3 and 4.

**Figure 3:**
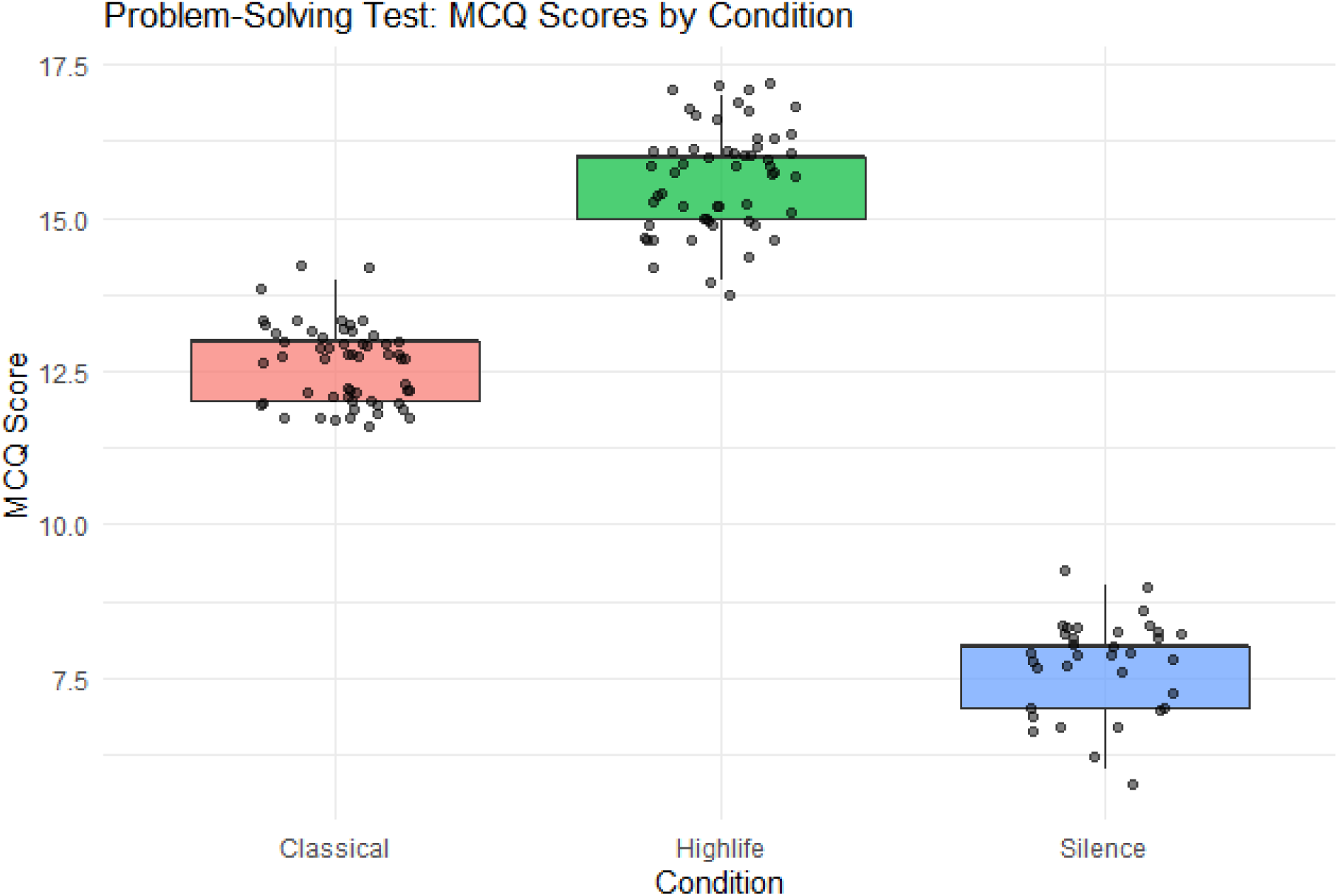
Problem-Solving Accuracy (MCQ Scores) by Auditory Condition.

**Figure 4:**
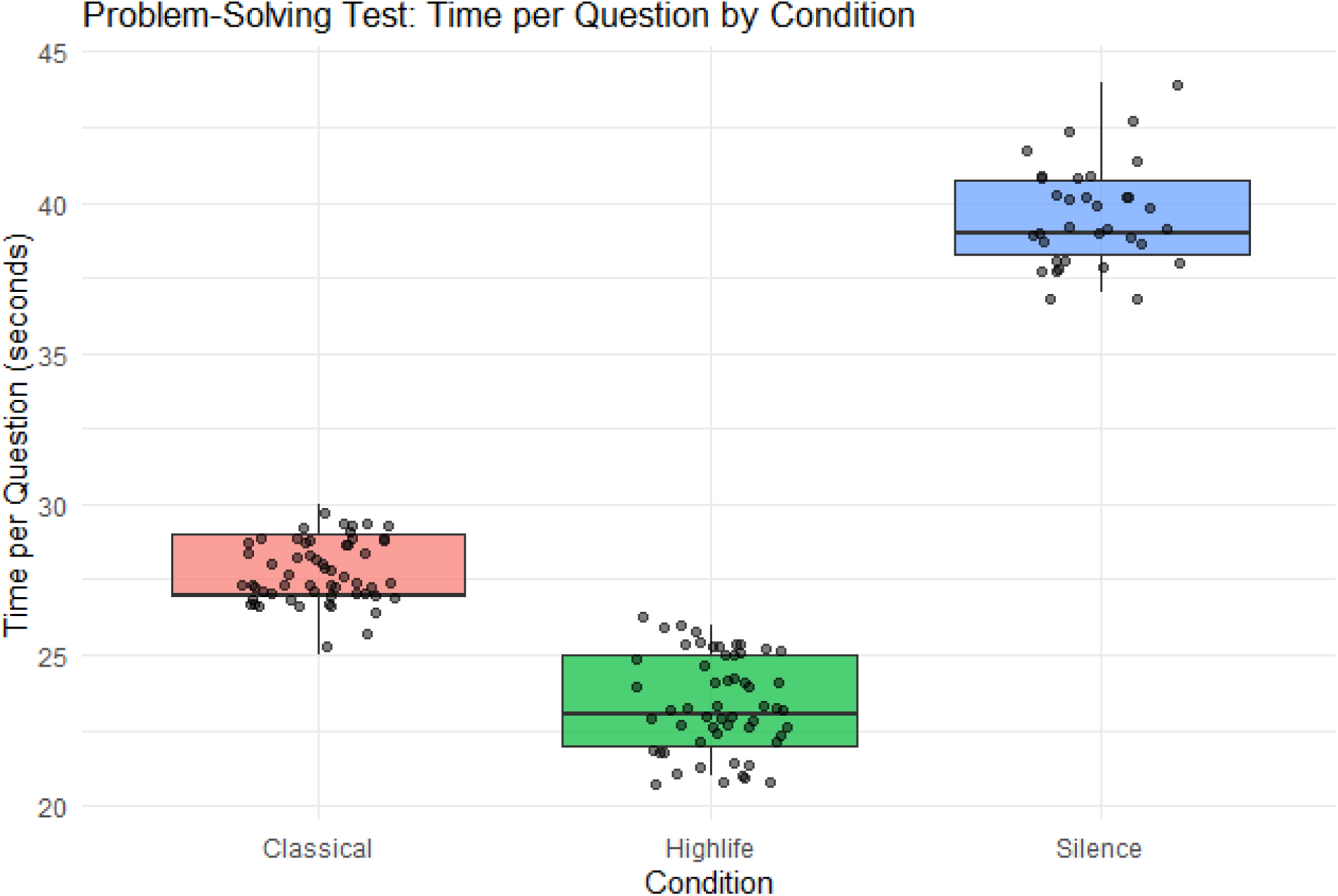
Problem-Solving Speed (Seconds per Question) by Auditory Condition.

**Figure 5:**
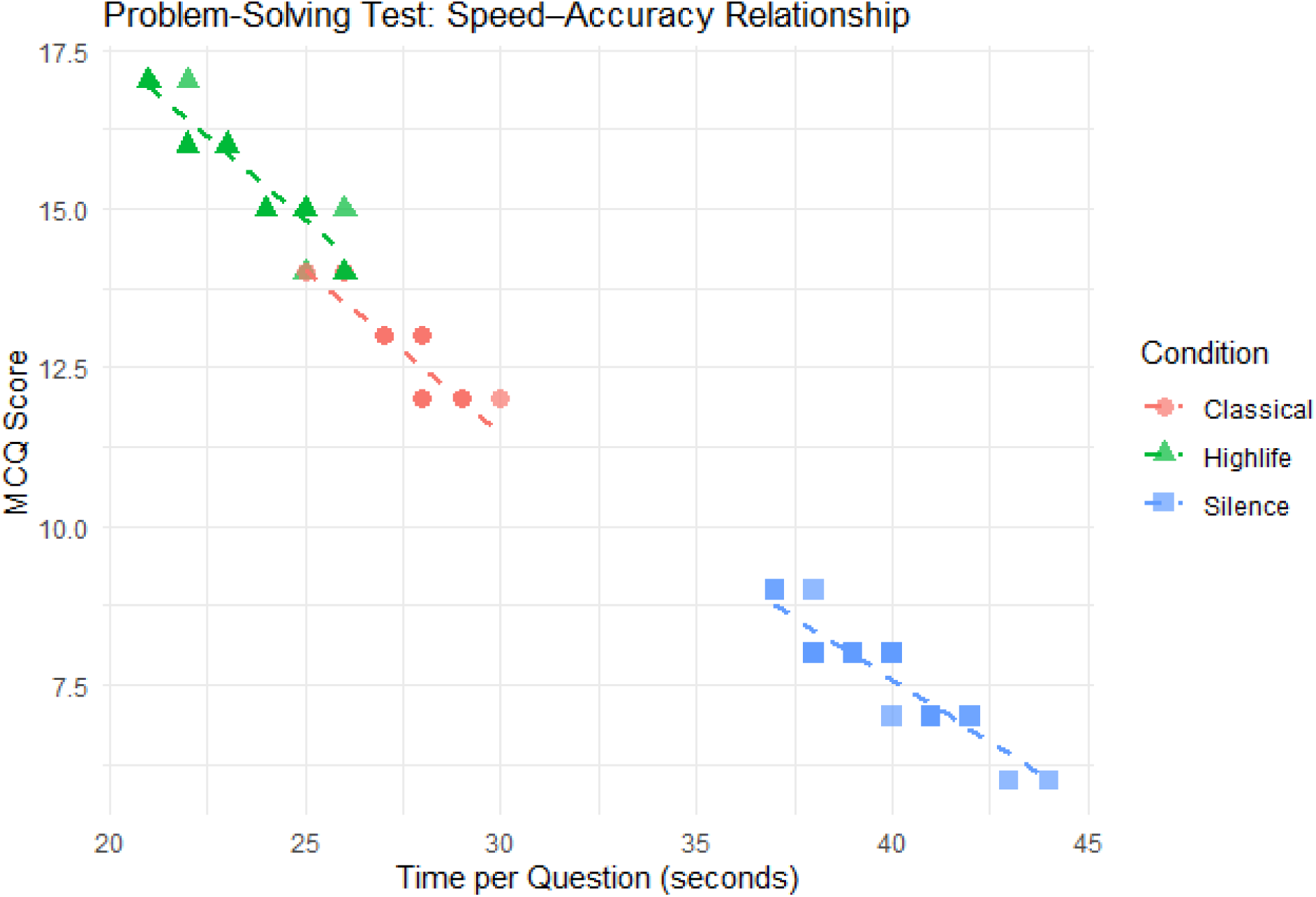
Speed-Accuracy Relationship by Auditory Condition.

**Table 6:** Problem-Solving Accuracy and Speed by Auditory Condition.

| Condition | n | Accuracy (Mean ± SD) | Speed (seconds/question) (Mean ± SD) |
| --- | --- | --- | --- |
| **Igbo Highlife** | 56 | 15.7 ± 0.855 | 23.4 ± 1.54 |
| **Western Classical** | 57 | 12.6 ± 0.587 | 27.7 ± 1.03 |
| **Silence** | 34 | 7.74 ± 0.710 | 39.6 ± 1.63 |

#### Multivariate Analysis

A one-way multivariate analysis of variance was conducted to examine the combined effect of auditory condition on problem-solving accuracy and speed. The results revealed a significant multivariate main effect (Pillai’s Trace = 1.656, F(4,288) = 346.72, p < 0.001), indicating that auditory condition significantly influenced the composite of accuracy and speed outcomes (Table 7).

**Table 7:** MANOVA Results for Problem-Solving Accuracy and Speed.

| Source | Pillai's Trace | Approx. F | Hypothesis df | Error df | p |
| --- | --- | --- | --- | --- | --- |
| **Condition** | 1.656 | 346.72 | 4 | 288 | <0.001 |

#### Univariate Analyses

Follow-up univariate ANOVAs demonstrated significant effects of condition on both accuracy (F(2,144) = 1263, p < 0.001, η²p = 0.95) and speed (F(2,144) = 1482, p < 0.001, η²p = 0.95) (Tables 8 and 9).

**Table 8:** One-Way ANOVA for Problem-Solving Accuracy by Condition.

| Source | df | Sum of Squares | Mean Square | F | p | $\eta^2p$ |
| --- | --- | --- | --- | --- | --- | --- |
| **Condition** | 2 | 1334.9 | 667.4 | 1263 | <0.001 | 0.95 |
| **Residuals** | 144 | 76.1 | 0.5 |  |  |  |

**Table 9:** One-Way ANOVA for Problem-Solving Speed by Condition.

| Source | df | Sum of Squares | Mean Square | F | P | $\eta^2p$ |
| --- | --- | --- | --- | --- | --- | --- |
| **Condition** | 2 | 5218.3 | 2609.1 | 1482 | <0.001 | 0.95 |
| **Residuals** | 144 | 253.6 | 1.76 |  |  |  |

Tukey’s HSD post-hoc comparisons for accuracy (Table 10) revealed that all pairwise differences were significant (p < 0.001). The Igbo Highlife condition significantly outperformed both Western Classical (mean difference = 3.05, 95% CI [2.72, 3.37]) and Silence (mean difference = 7.94, 95% CI [7.57, 8.32]). Western Classical also significantly outperformed Silence (mean difference = 4.90, 95% CI [4.52, 5.27]).

**Table 10:** Tukey’s HSD Post-Hoc Comparisons for Problem-Solving Accuracy.

| Comparison | Mean Difference | 95% CI Lower | 95% CI Upper | p |
| --- | --- | --- | --- | --- |
| **Highlife – Classical** | 3.05 | 2.72 | 3.37 | <0.001 |
| **Silence – Classical** | -4.90 | -5.27 | -4.52 | <0.001 |
| **Silence – Highlife** | -7.94 | -8.32 | -7.57 | <0.001 |

### Relationship Between Cultural Familiarity and Cognitive Performance

#### Descriptive Statistics by Familiarity Level

Participants were categorized based on self-reported familiarity with Igbo Highlife music. Table 11 presents problem-solving performance stratified by familiarity level, revealing a gradient of improvement with increasing familiarity.

**Table 11:** Problem-Solving Performance by Highlife Familiarity Level.

| Familiarity Level | n | Accuracy (Mean ± SD) | Speed (seconds) (Mean ± SD) |
| --- | --- | --- | --- |
| **Not at all familiar** | 15 | 10.7 ± 3.24 | 32.9 ± 7.33 |
| **Slightly familiar** | 20 | 12.0 ± 3.26 | 30.4 ± 6.85 |
| **Moderately familiar** | 32 | 12.6 ± 2.80 | 28.6 ± 5.75 |
| **Very familiar** | 80 | 13.2 ± 3.04 | 27.7 ± 6.08 |

#### Correlation Analyses

Spearman’s rank correlation analyses (Table 12) revealed significant relationships between cultural familiarity and cognitive performance. Familiarity with Igbo Highlife was positively correlated with problem-solving accuracy (ρ = 0.268, p = 0.001) and negatively correlated with completion time (ρ = -0.263, p = 0.001), indicating that greater familiarity was associated with both higher accuracy and faster processing.

**Table 12:** Correlations Between Highlife Familiarity and Cognitive Performance.

| Variable Pair | $\rho$ | p |
| --- | --- | --- |
| **Familiarity – Accuracy (MCQ Score)** | 0.268 | 0.001 |
| **Familiarity – Speed (Time per Question)** | -0.263 | 0.001 |

#### Two-Way ANOVA: Familiarity Group × Condition

To further examine the role of cultural familiarity, participants were dichotomized into High Familiarity (n = 72) and Low-to-Moderate Familiarity (n = 75) groups. A two-way ANOVA (Table 13) examined the effects of Familiarity Group and Auditory Condition on cognitive performance.

**Table 13:** Two-Way ANOVA for Familiarity Group and Condition on Performance.

| Source | df | Sum Sq | Mean Sq | F | p |
| --- | --- | --- | --- | --- | --- |
| **Familiarity Group** | 1 | 37.0 | 37.0 | 67.02 | <0.001 |
| **Condition** | 2 | 1294.5 | 647.2 | 1172.64 | <0.001 |
| **Familiarity Group × Condition** | 2 | 1.7 | 0.8 | 1.54 | 0.219 |
| **Residuals** | 141 | 77.8 | 0.6 |  |  |

Significant main effects were observed for both Familiarity Group (F(1,141) = 67.02, p < 0.001) and Condition (F(2,141) = 1172.64, p < 0.001). The interaction between Familiarity Group and Condition was not significant (F(2,141) = 1.54, p = 0.219), indicating that the pattern of benefit across conditions was similar for both familiarity groups, though the High Familiarity group demonstrated consistently higher baseline performance.

Detailed pairwise comparisons (Table 14) confirmed that within the Highlife condition, participants with High Familiarity significantly outperformed those with Low-to-Moderate Familiarity (mean difference = 1.21, p < 0.001), demonstrating an additive benefit of cultural familiarity.

**Table 14:**
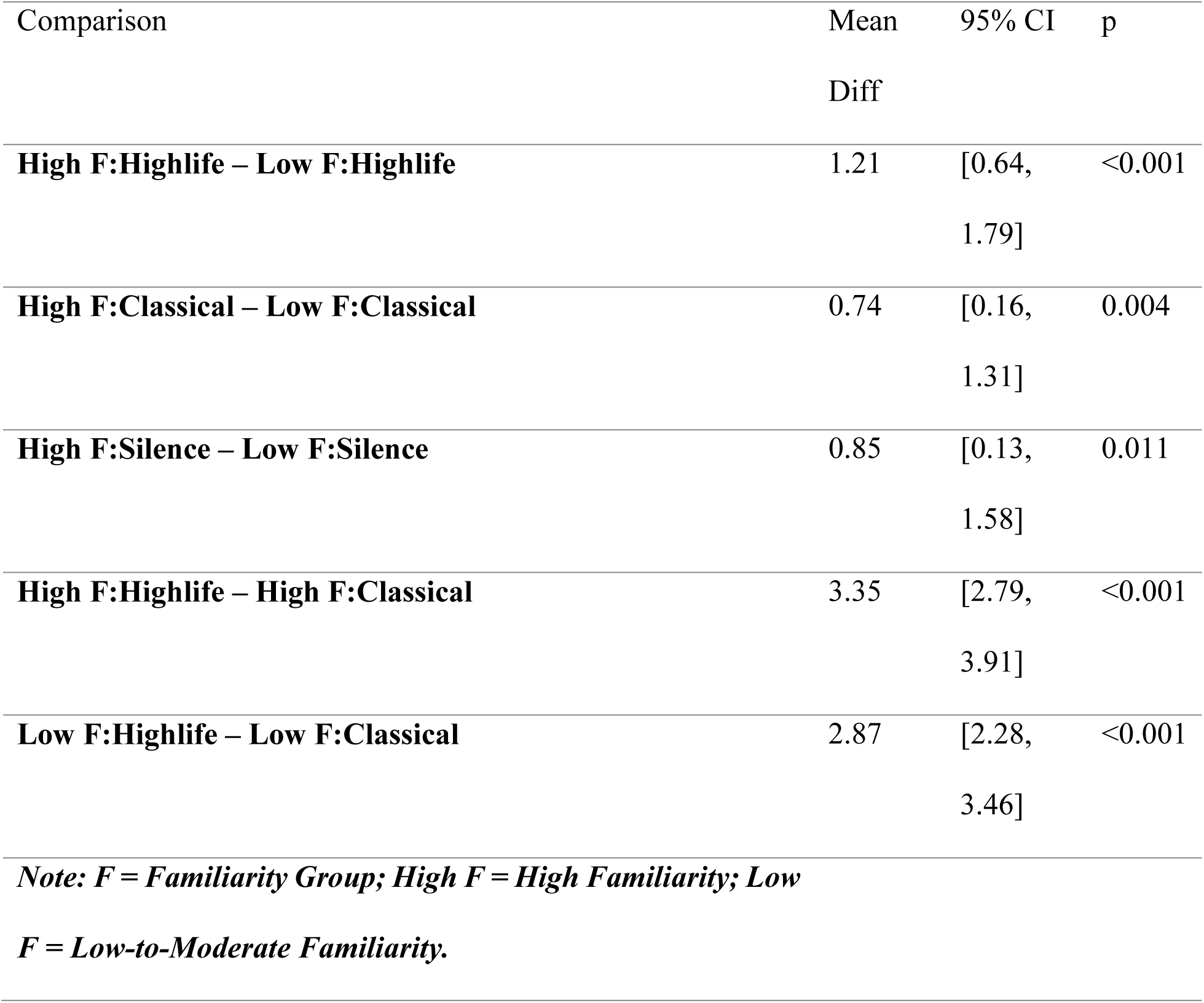
Selected Tukey’s HSD Pairwise Comparisons for Familiarity Group × Condition.

| Comparison | Mean<br>Diff | 95% CI | p |
| --- | --- | --- | --- |
| **High F:Highlife – Low F:Highlife** | 1.21 | [0.64,<br>1.79] | <0.001 |
| **High F:Classical – Low F:Classical** | 0.74 | [0.16,<br>1.31] | 0.004 |
| **High F:Silence – Low F:Silence** | 0.85 | [0.13,<br>1.58] | 0.011 |
| **High F:Highlife – High F:Classical** | 3.35 | [2.79,<br>3.91] | <0.001 |
| **Low F:Highlife – Low F:Classical** | 2.87 | [2.28,<br>3.46] | <0.001 |
| *Note: F = Familiarity Group; High F = High Familiarity; Low F = Low-to-Moderate Familiarity.* |  |  |  |

## DISCUSSION

This randomized controlled trial investigated the effects of culturally familiar background music (Igbo Highlife) on cognitive performance among Nigerian medical students, comparing outcomes with Western classical music and silence. The findings provide robust evidence that background music, particularly when culturally salient, significantly enhances both memory recall and clinical problem-solving performance.

### Effect of Igbo Highlife on Memory Recall

The first major finding was that participants exposed to Igbo Highlife demonstrated superior memory recall compared to both Western classical music and silence. The mean recall score in the Highlife condition (16.7/20) was substantially higher than in the classical (13.7/20) and silence (10.2/20) conditions, with all pairwise comparisons reaching statistical significance (p < 0.001). This pattern confirms our first hypothesis.

These findings align with theoretical frameworks proposing that rhythmic predictability and emotional engagement optimize arousal and facilitate neural entrainment. The superior performance of Highlife over classical music specifically suggests that cultural familiarity reduces the cognitive load required to process auditory stimuli, thereby freeing attentional resources for the primary memory task [5]. The silence condition, lacking this beneficial modulation of arousal and potentially permitting environmental distraction, resulted in the poorest performance.

This finding extends the music-cognition literature beyond the traditional Eurocentric focus of the “Mozart Effect” [2] by demonstrating that culturally indigenous music can produce cognitive enhancements comparable to—and indeed exceeding—those observed with Western classical stimuli. The effect size (η²p = 0.91) indicates that auditory condition explained the vast majority of variance in memory performance, underscoring the practical significance of this manipulation.

### Effect of Igbo Highlife on Problem-Solving

The second major finding addressed our second hypothesis: Igbo Highlife significantly enhanced both the accuracy and speed of clinical problem-solving. Participants in the Highlife condition achieved the highest accuracy (15.7/20) while simultaneously completing tasks most quickly (23.4 seconds/question). The MANOVA confirmed that auditory condition significantly influenced the composite of accuracy and speed (p < 0.001), with Highlife demonstrating the strongest combined effect.

This pattern aligns with cognitive-affective theories of learning, which posit that familiar, non-verbal stimuli consume fewer attentional resources, enabling more efficient allocation of cognitive capacity to primary tasks. The significantly worse performance in the silence condition contradicts the common student belief that silence is optimal for complex cognitive work, suggesting that appropriate background music may serve as a facilitative stimulus by maintaining optimal arousal and reducing mind-wandering.

The graded pattern of performance (Highlife > Classical > Silence) is particularly noteworthy. While classical music conferred significant benefits relative to silence, its impact was less pronounced than Highlife, highlighting the importance of cultural relevance in music-mediated cognitive enhancement. This finding has practical implications for medical education in non-Western settings, suggesting that locally relevant musical stimuli may be more effective than imported Western genres.

### The Moderating Role of Cultural Familiarity

The third major finding addressed our third hypothesis: cultural familiarity with Highlife significantly correlated with enhanced cognitive outcomes. The positive correlation between familiarity and accuracy (ρ = 0.268, p = 0.001) and negative correlation with completion time (ρ = -0.263, p = 0.001) indicate that greater familiarity was associated with both higher accuracy and faster processing.

The two-way ANOVA revealed significant main effects of both familiarity group and auditory condition, with no significant interaction. This pattern indicates that while all participants benefited from the Highlife stimulus, those with greater prior familiarity derived an additional performance advantage. The additive nature of this benefit—where familiarity elevated baseline performance across all conditions and amplified the advantage within the Highlife condition—provides strong support for the enculturation hypothesis [6].

These findings are consistent with prior evidence that familiar stimuli increase neural efficiency and engagement. The observation that even within the silence condition, highly familiar participants performed better (mean difference = 0.85, p = 0.011) suggests that cultural familiarity may confer broader cognitive advantages beyond music-specific effects, possibly through enhanced metacognitive strategies or greater engagement with culturally congruent testing contexts.

### Integration with Theoretical Frameworks

The results of this study can be integrated with three complementary theoretical frameworks. First, consistent with the Arousal-Mood Hypothesis [4], the emotionally engaging nature of culturally familiar Highlife may have optimized affective states for cognitive processing. Second, in accordance with Cognitive Load Theory [5], the reduced processing demands of familiar auditory stimuli may have freed working memory resources for primary task performance. Third, supporting the Theory of Enculturation [6], prolonged exposure to the rhythmic and melodic syntax of Highlife likely enabled more efficient neural processing among familiar listeners.

The absence of a significant interaction between familiarity and condition suggests that the cognitive benefits of Highlife are not exclusively dependent on prior familiarity, though familiarity provides an additive advantage. This may reflect intrinsic properties of the music itself—such as its rhythmic complexity or polyphonic structure—that facilitate neural entrainment even among unfamiliar listeners.

### Strengths and Limitations

This study possesses several methodological strengths, including its randomized controlled design, adequate sample size determined by a priori power analysis, blinding of the analyst, and use of validated outcome measures. The inclusion of both immediate and delayed components, as well as multiple cognitive domains (memory and problem-solving), enhances the generalizability of findings.

Several limitations should be acknowledged. First, the study was limited to a single genre of Nigerian music (Igbo Highlife) and did not explore other indigenous musical forms. Second, the use of instrumental tracks may not capture the full effect of lyrical music, which could either enhance or impair cognitive performance depending on content and task demands. Third, the controlled laboratory setting may not fully generalize to real-world study environments with their characteristic distractions and variability. Fourth, the sample sizes across conditions were not evenly balanced, with the Silence group (n = 34) being smaller than the music groups, potentially limiting statistical power for some comparisons. Fifth, the delayed recall assessment was conducted at only one time point (48 hours), precluding examination of longer-term retention effects.

### Implications for Practice and Future Research

The findings of this study have several practical implications. For medical students in Nigeria and similar cultural contexts, incorporating familiar instrumental music into study routines may enhance learning efficiency. For educational institutions, these results support the integration of culturally relevant auditory stimuli into learning environments and wellness programs. More broadly, the findings challenge the universalist assumptions underlying much music-cognition research and suggest that educational interventions should be culturally tailored to maximize effectiveness.

Future research should address several questions arising from this study. First, replication studies should examine whether similar effects obtain with other Nigerian musical genres (e.g., Fuji, Juju, Apala) and in other cultural contexts. Second, neuroimaging studies could elucidate the neural mechanisms underlying the familiarity advantage observed here, potentially revealing differential activation patterns in auditory cortex and hippocampal regions. Third, longitudinal studies could examine whether the cognitive benefits of culturally familiar music persist over extended periods and translate to real-world academic outcomes such as examination performance. Fourth, studies manipulating the lyrical content of music could clarify the conditions under which lyrics facilitate versus impair cognitive processing.

## Conclusions

This study provides robust empirical evidence that background music, particularly culturally salient Igbo Highlife music, significantly enhances cognitive performance in medical students. The findings offer a nuanced answer to the long-debated “Mozart Effect,” suggesting that a “Highlife Effect” may be more powerful within its cultural context. Culturally familiar music consistently outperformed both Western classical music and silence in enhancing memory recall, problem-solving accuracy, and processing speed. The moderating role of cultural familiarity underscores the importance of considering cultural context in cognitive neuroscience research and educational practice. These outcomes suggest that integrating culturally relevant music into educational settings may represent an effective, non-invasive, and culturally congruent strategy for enhancing cognitive performance in medical education and beyond.

## Data Availability

All data produced in the present work are contained in the manuscript

## APPENDIX A: INFORMED CONSENT FORM

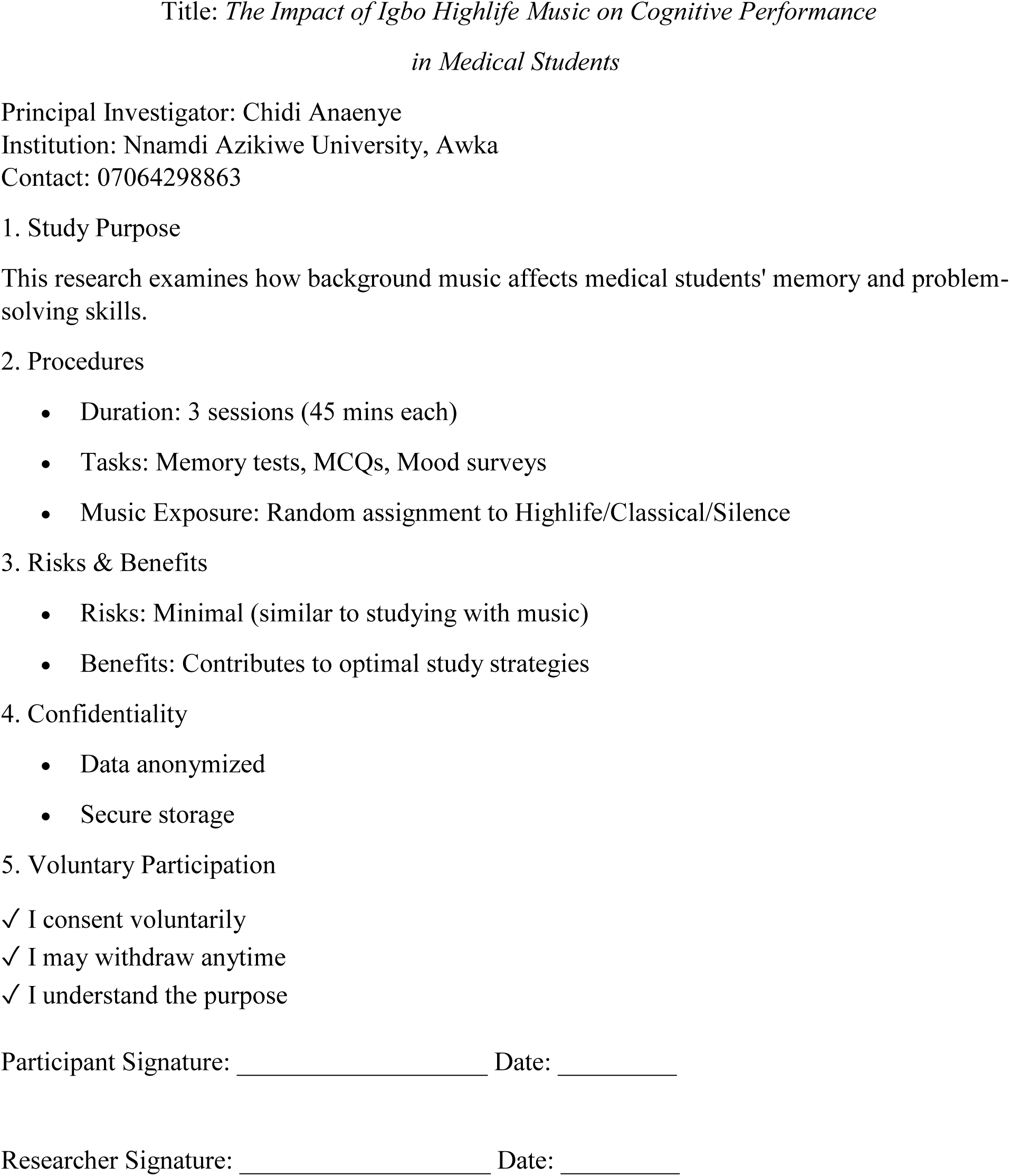

## APPENDIX B: DEMOGRAPHIC SURVEY

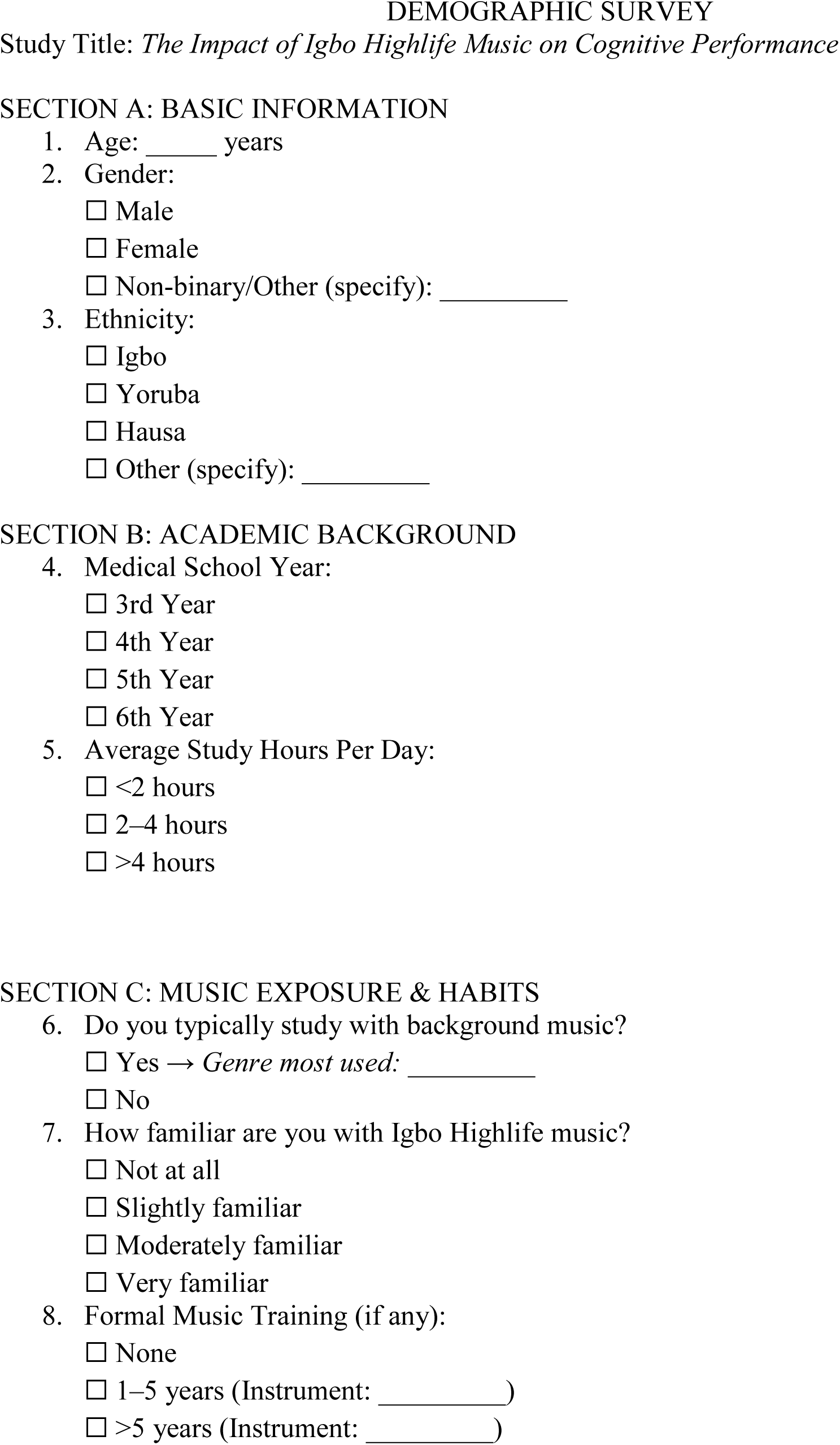
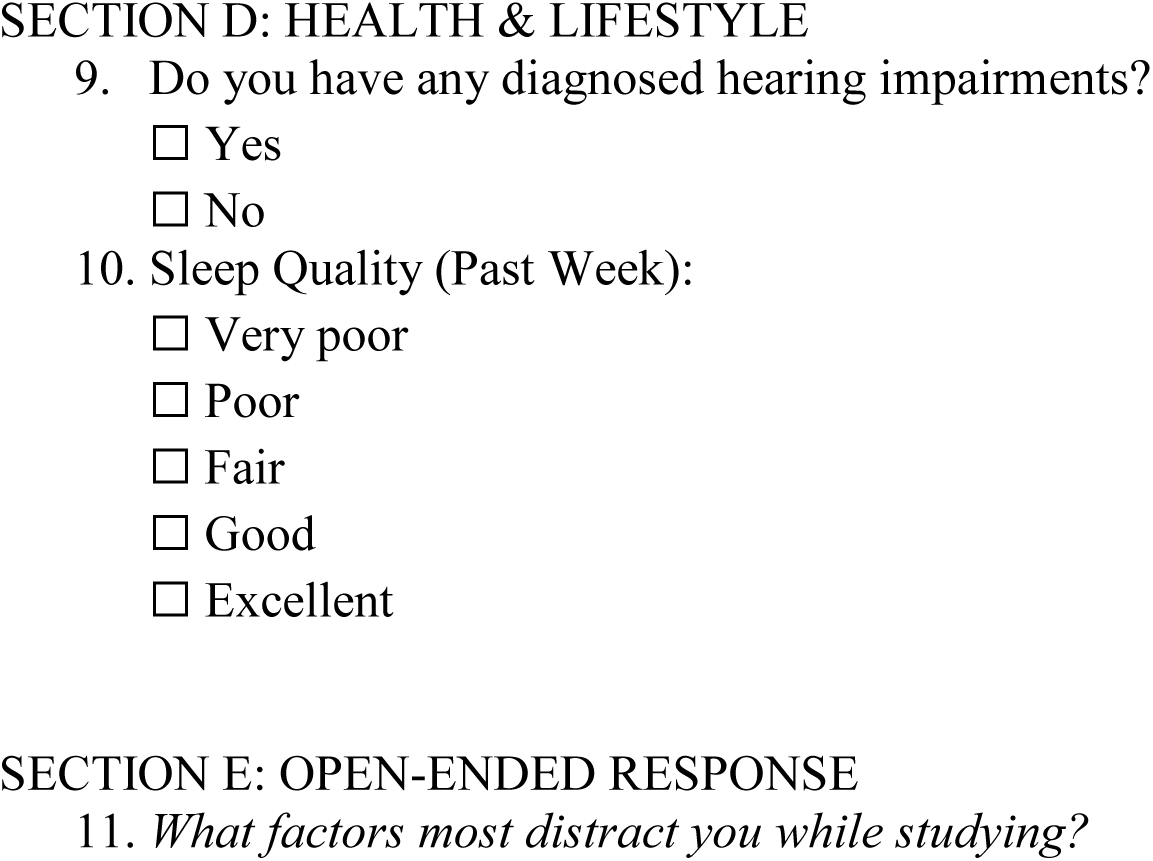

## APPENDIX C: BASELINE MOOD ASSESSMENT (PANAS)

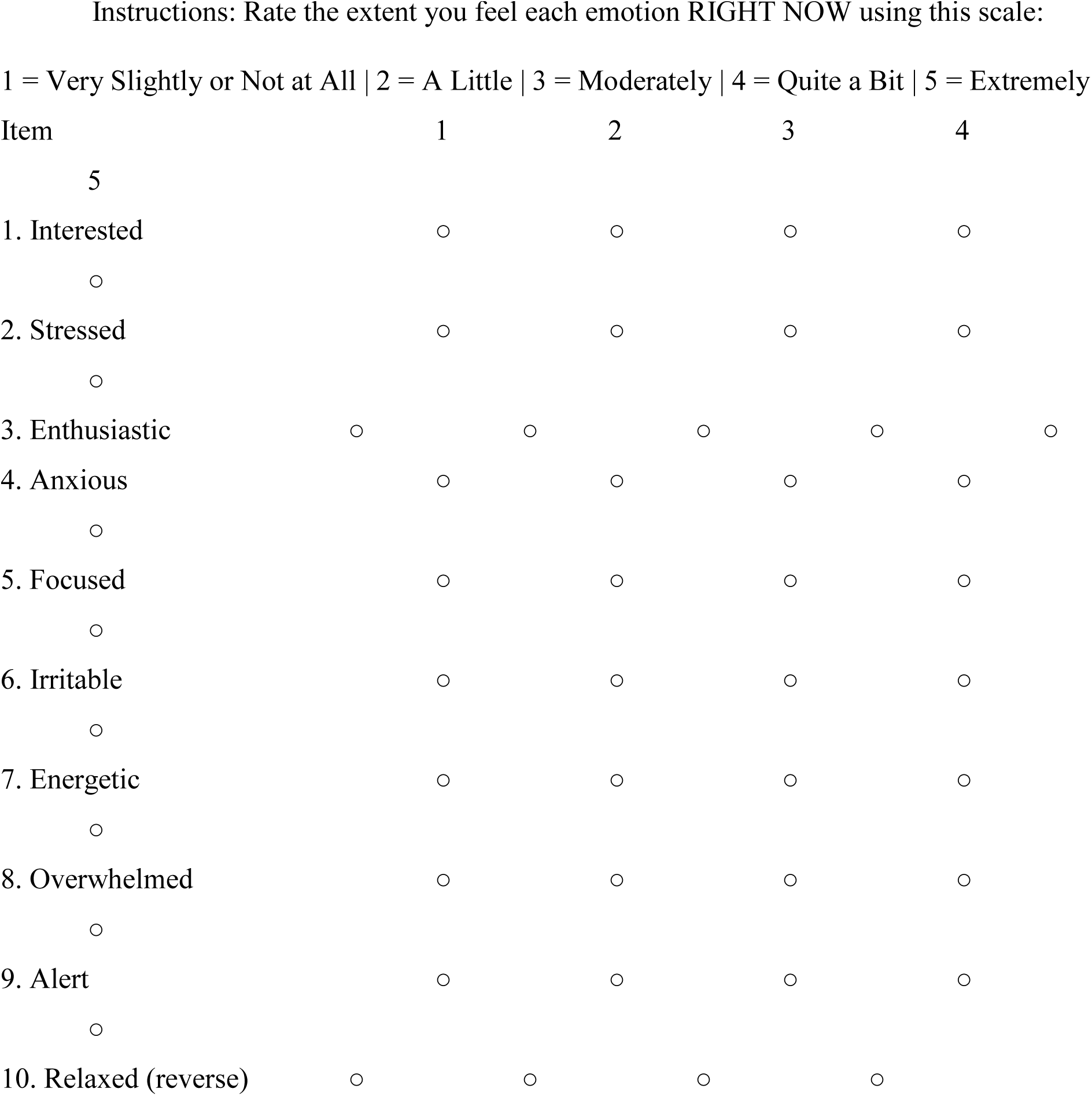

## APPENDIX D: MEMORY RECALL TEST

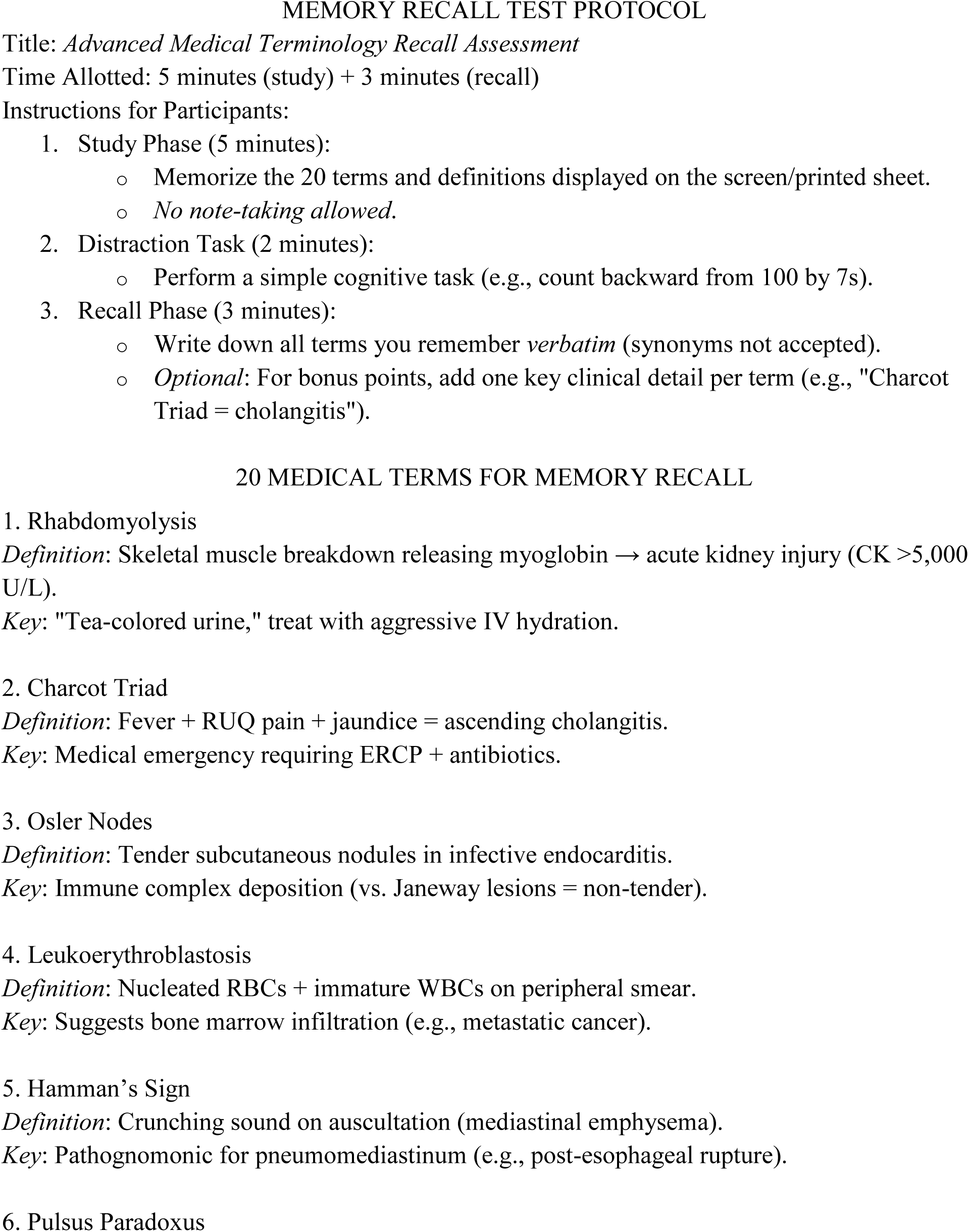
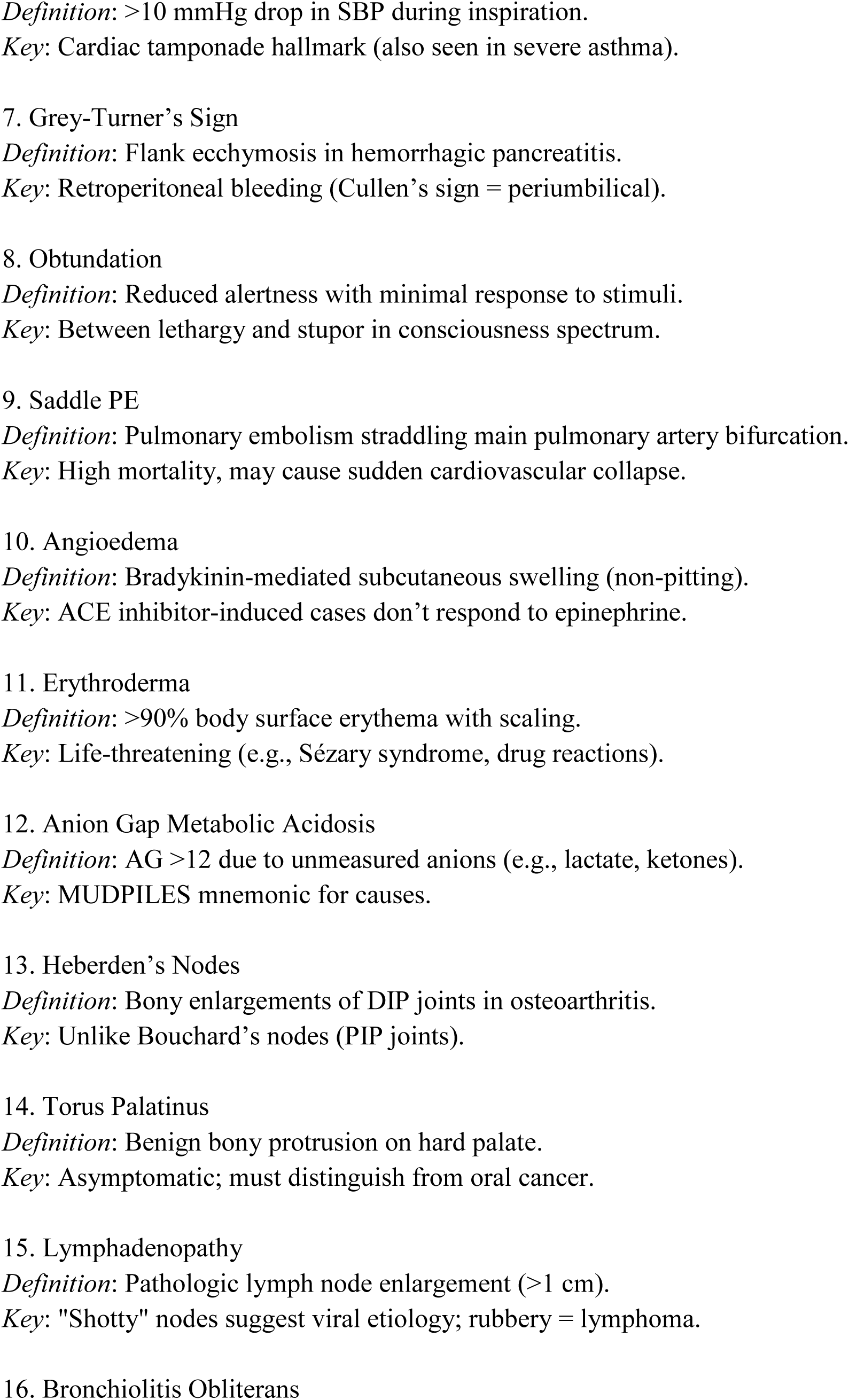
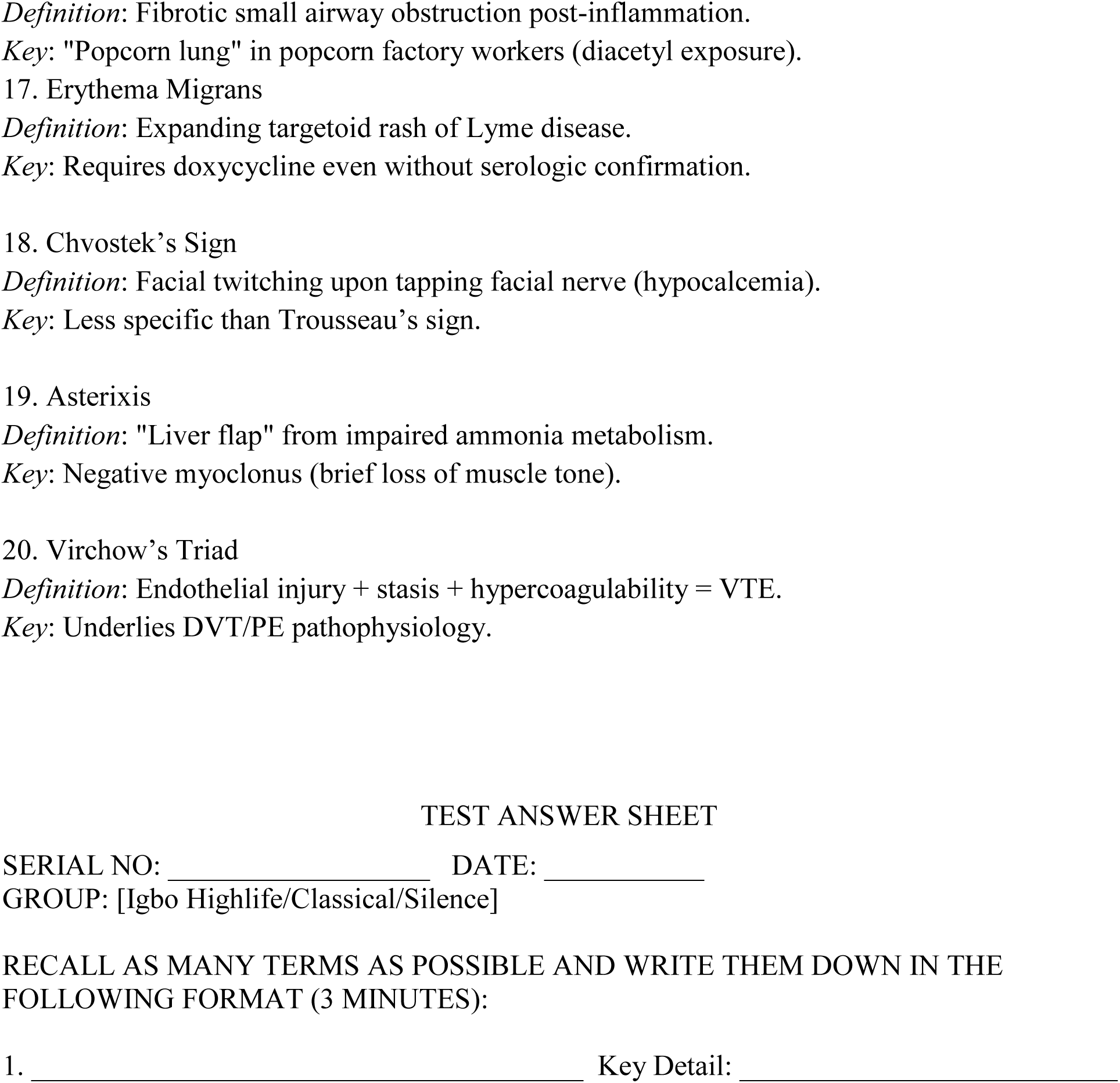

## APPENDIX E: PARALLEL MEMORY RECALL TEST

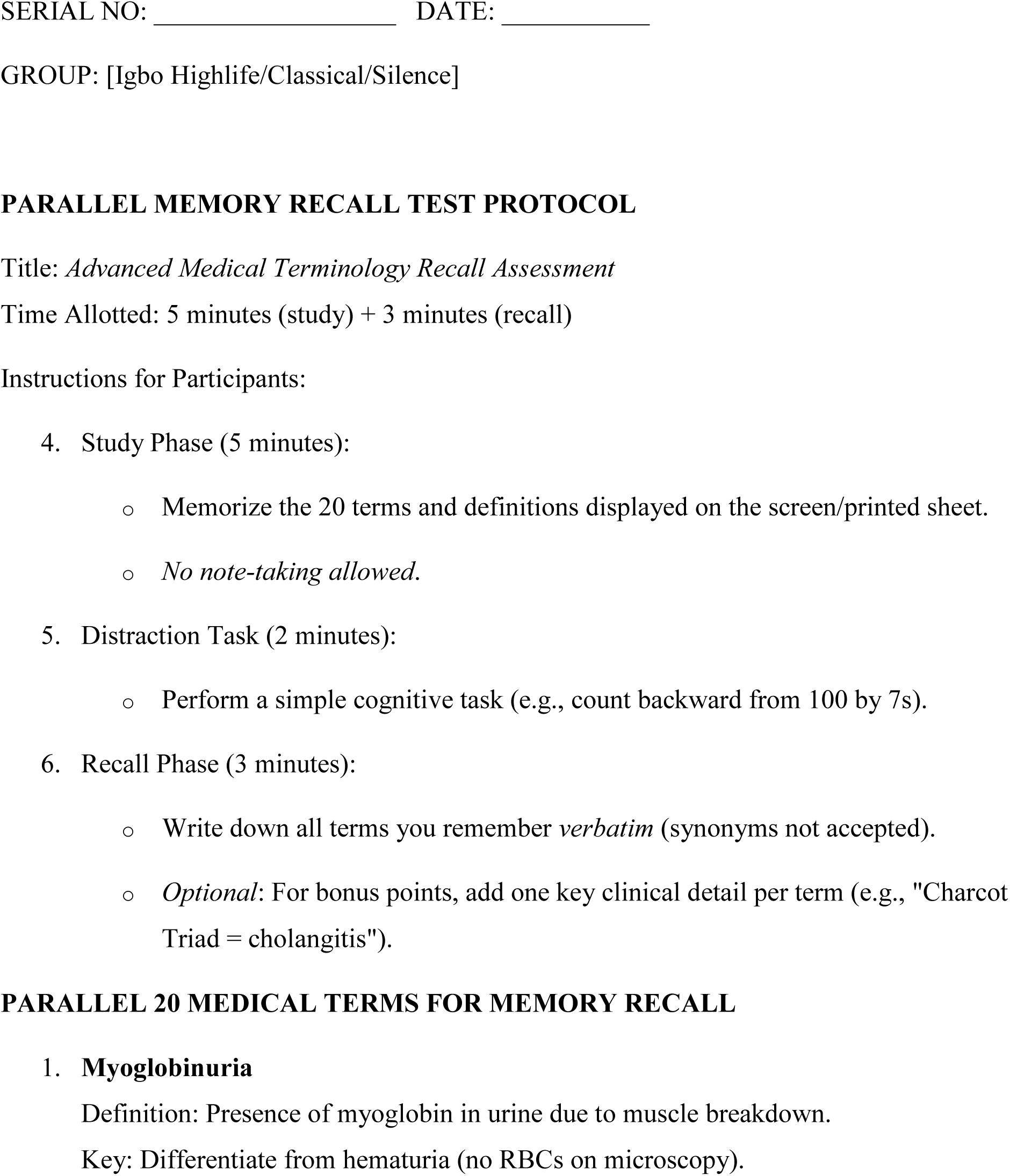
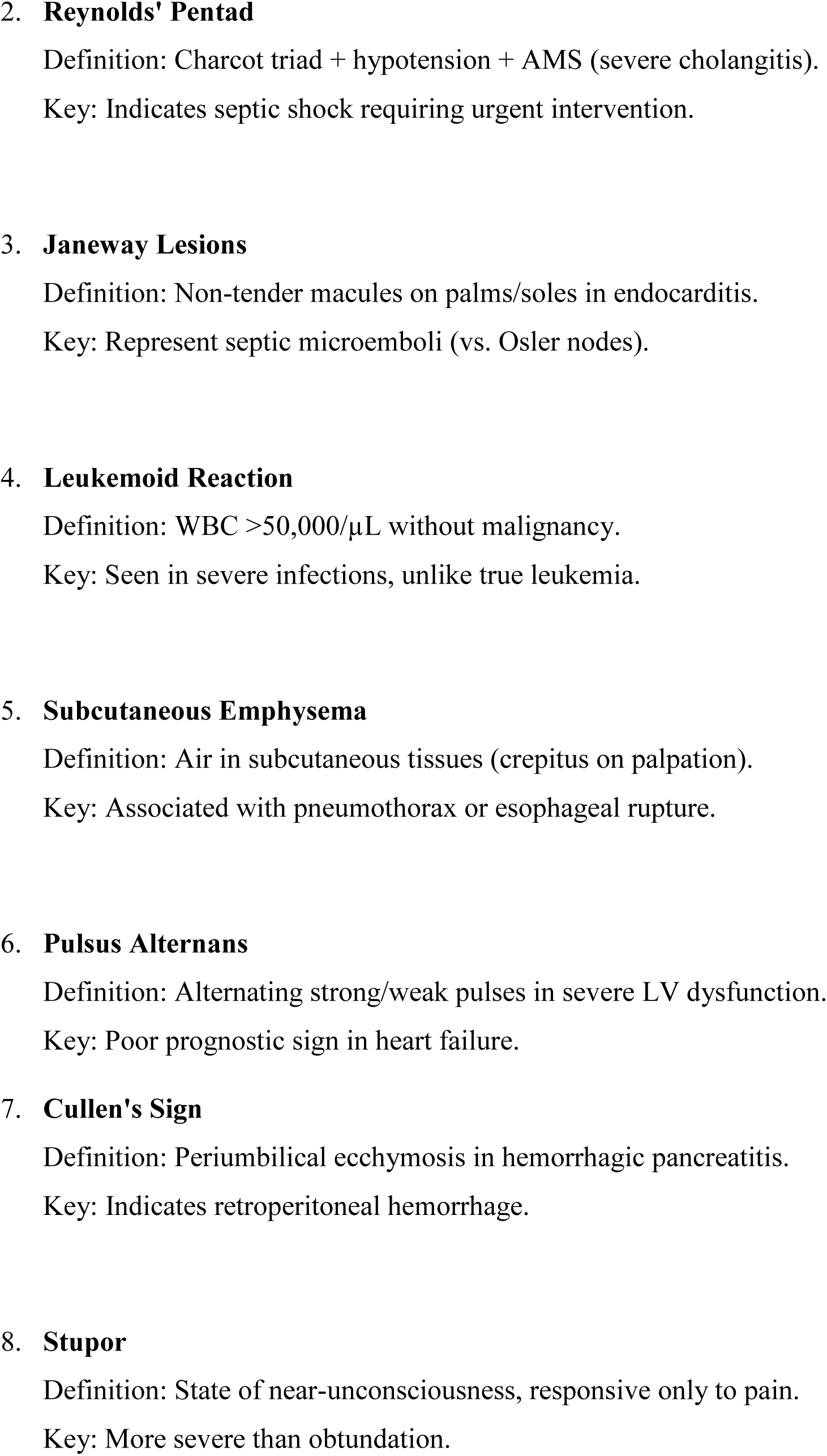
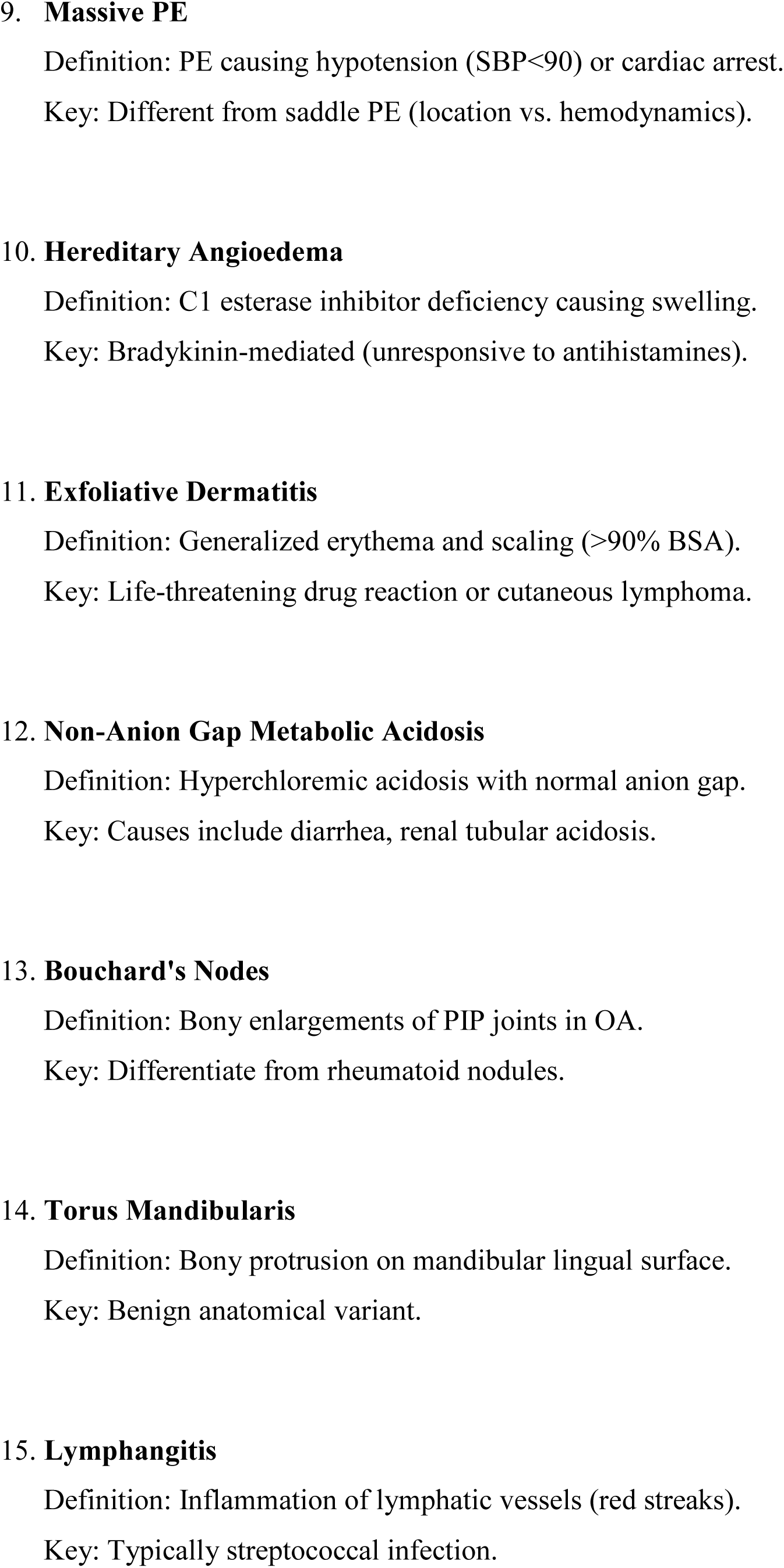
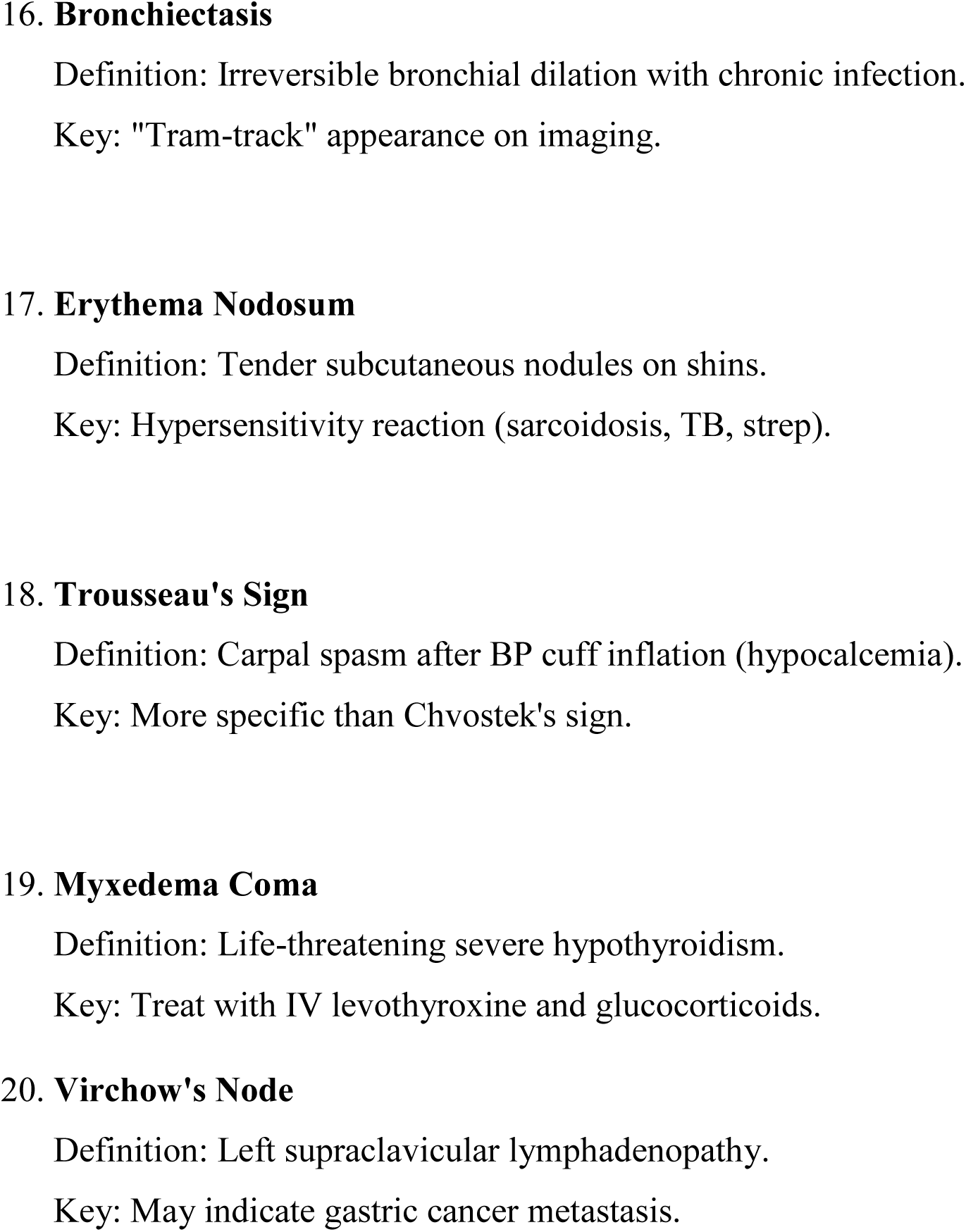

## APPENDIX F: PROBLEM-SOLVING MCQ (SAMPLE)

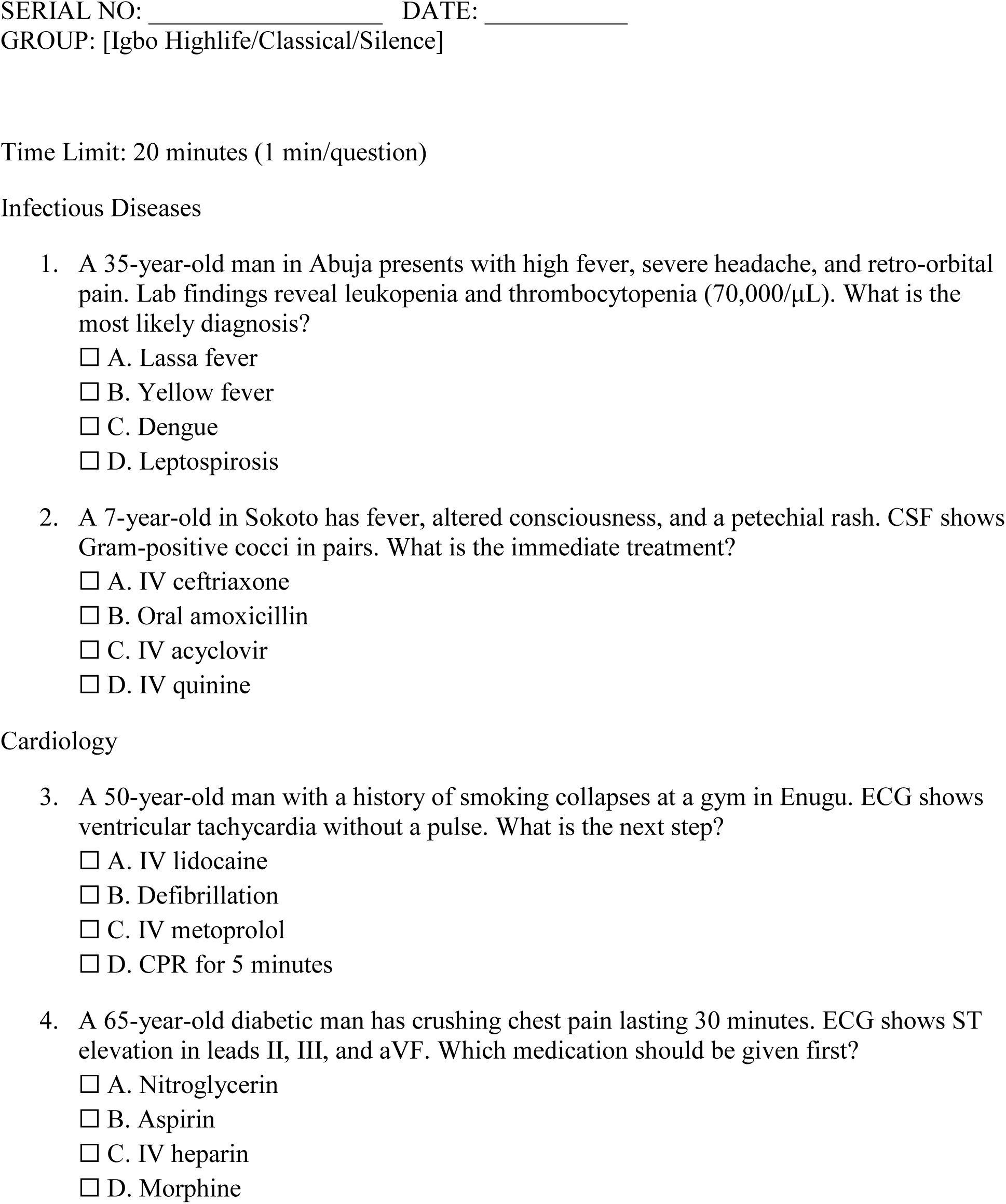
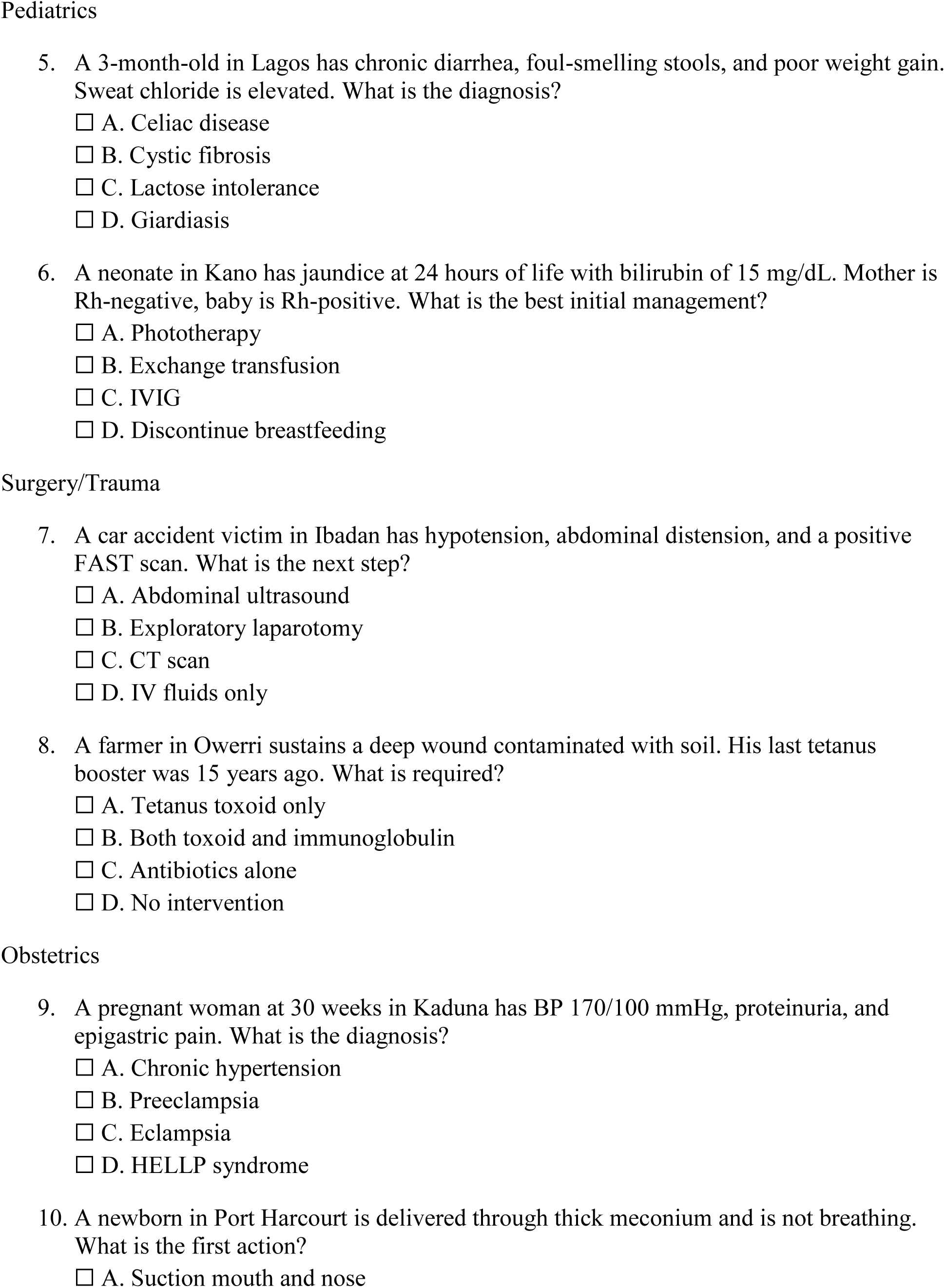
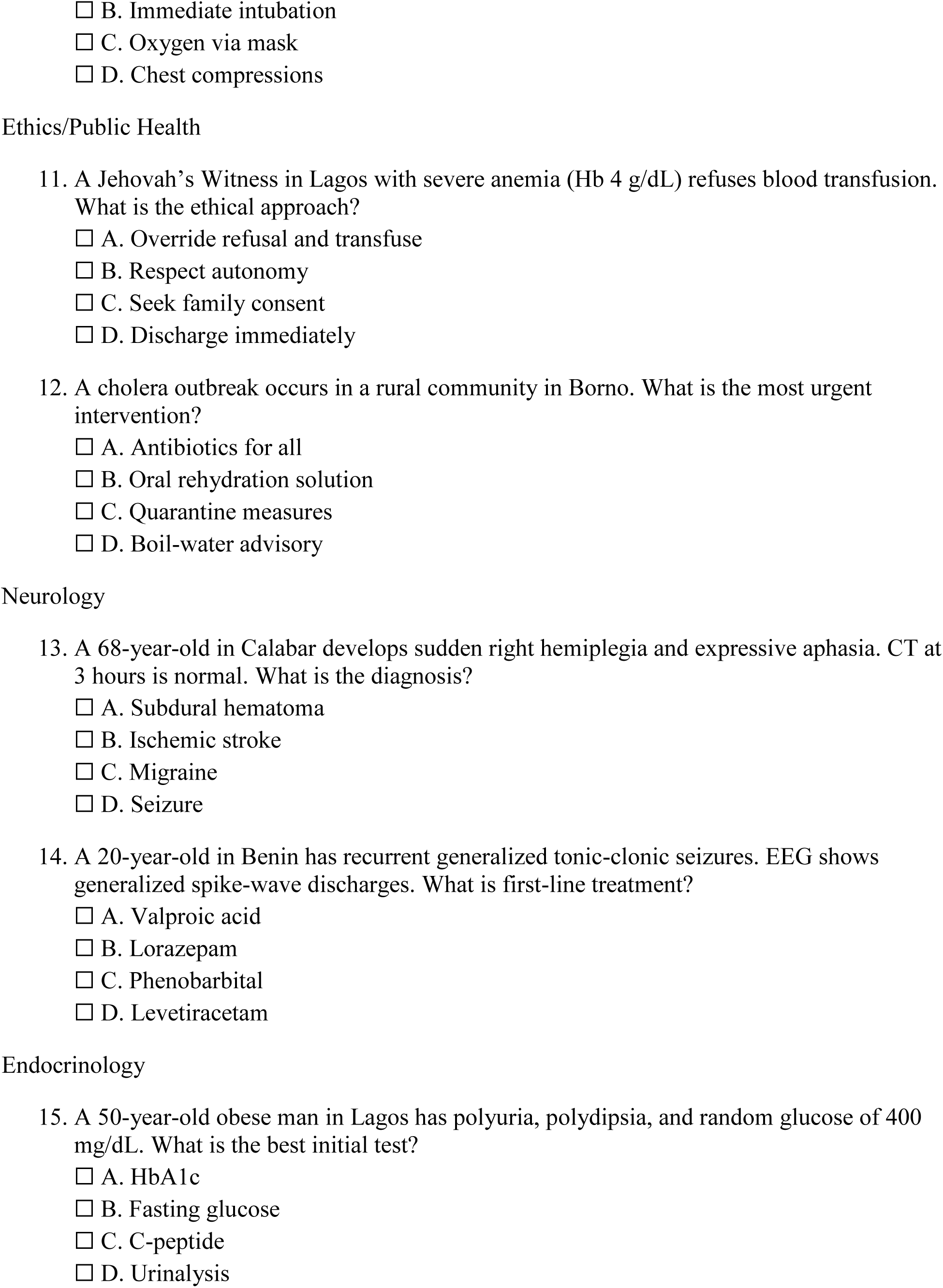
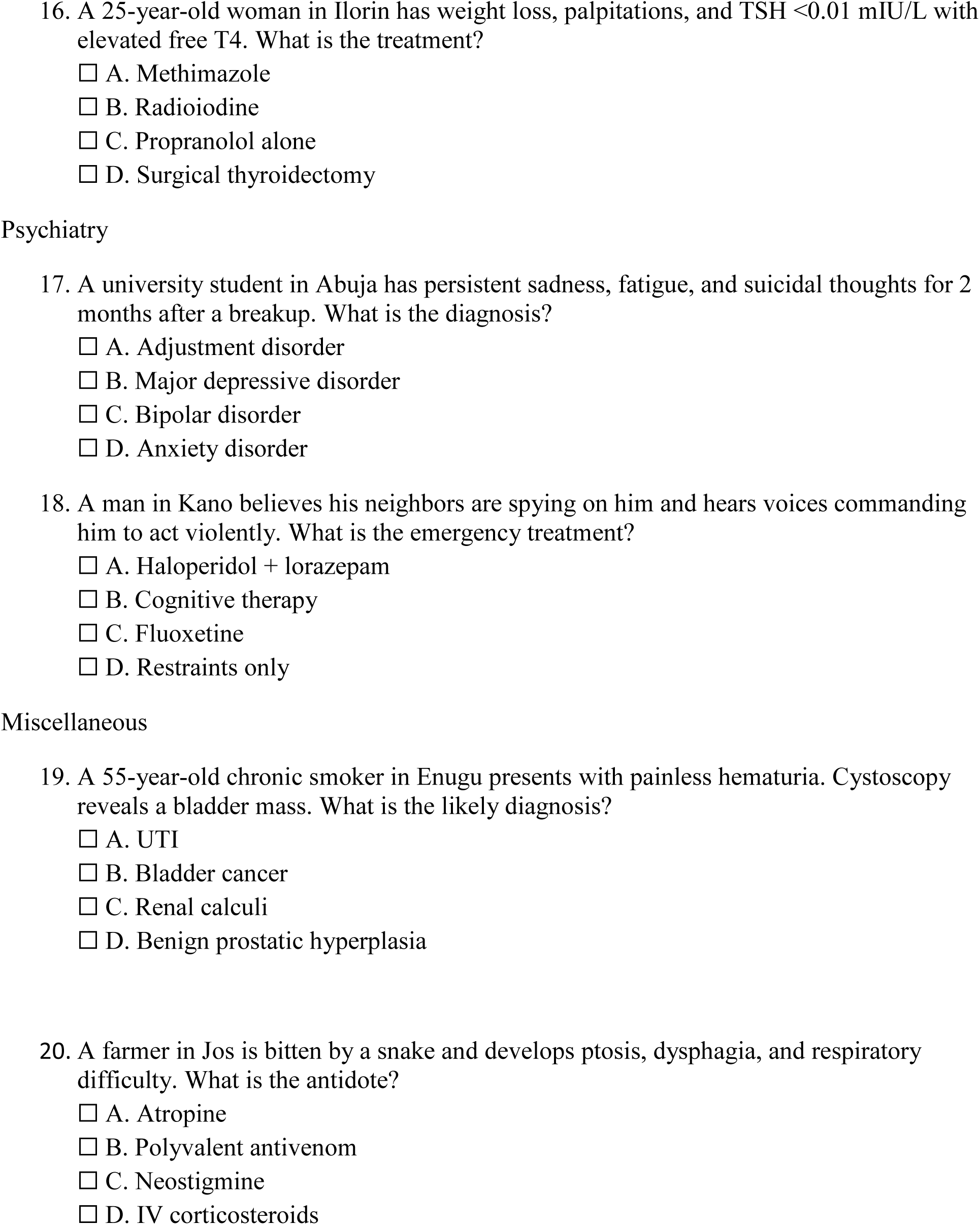

## APPENDIX G: PARALLEL TEST PROBLEM SOLVING ABILITY

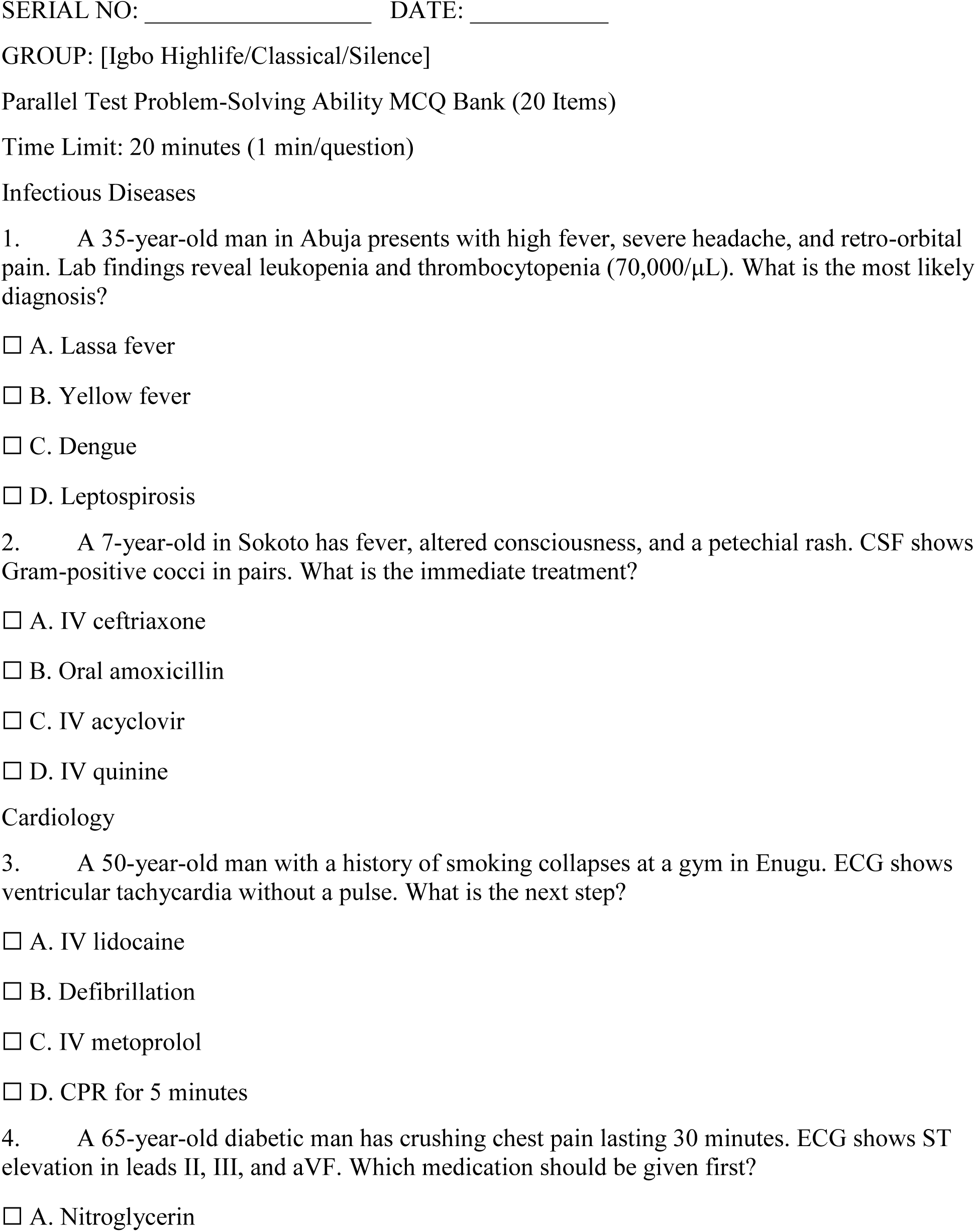
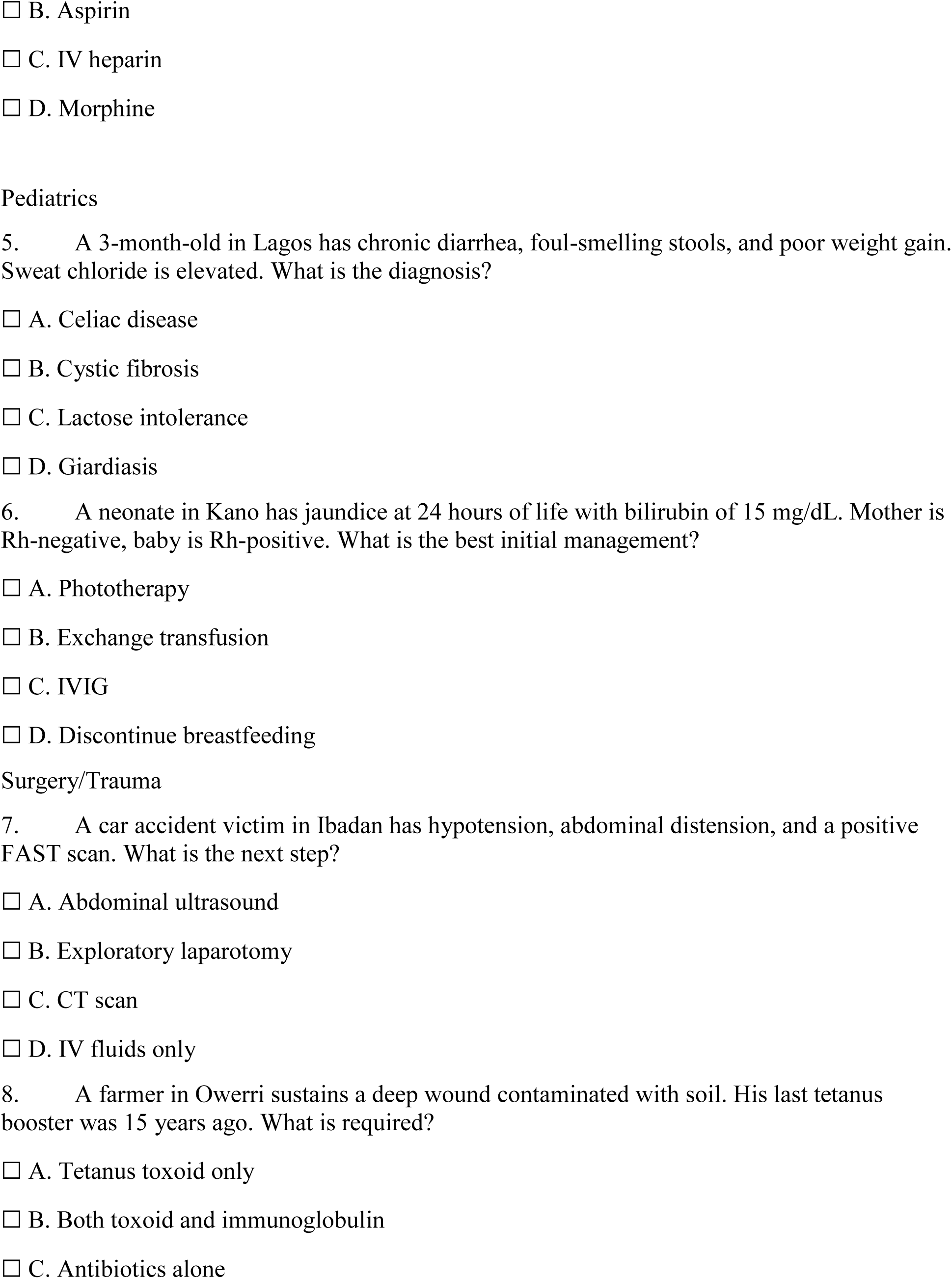
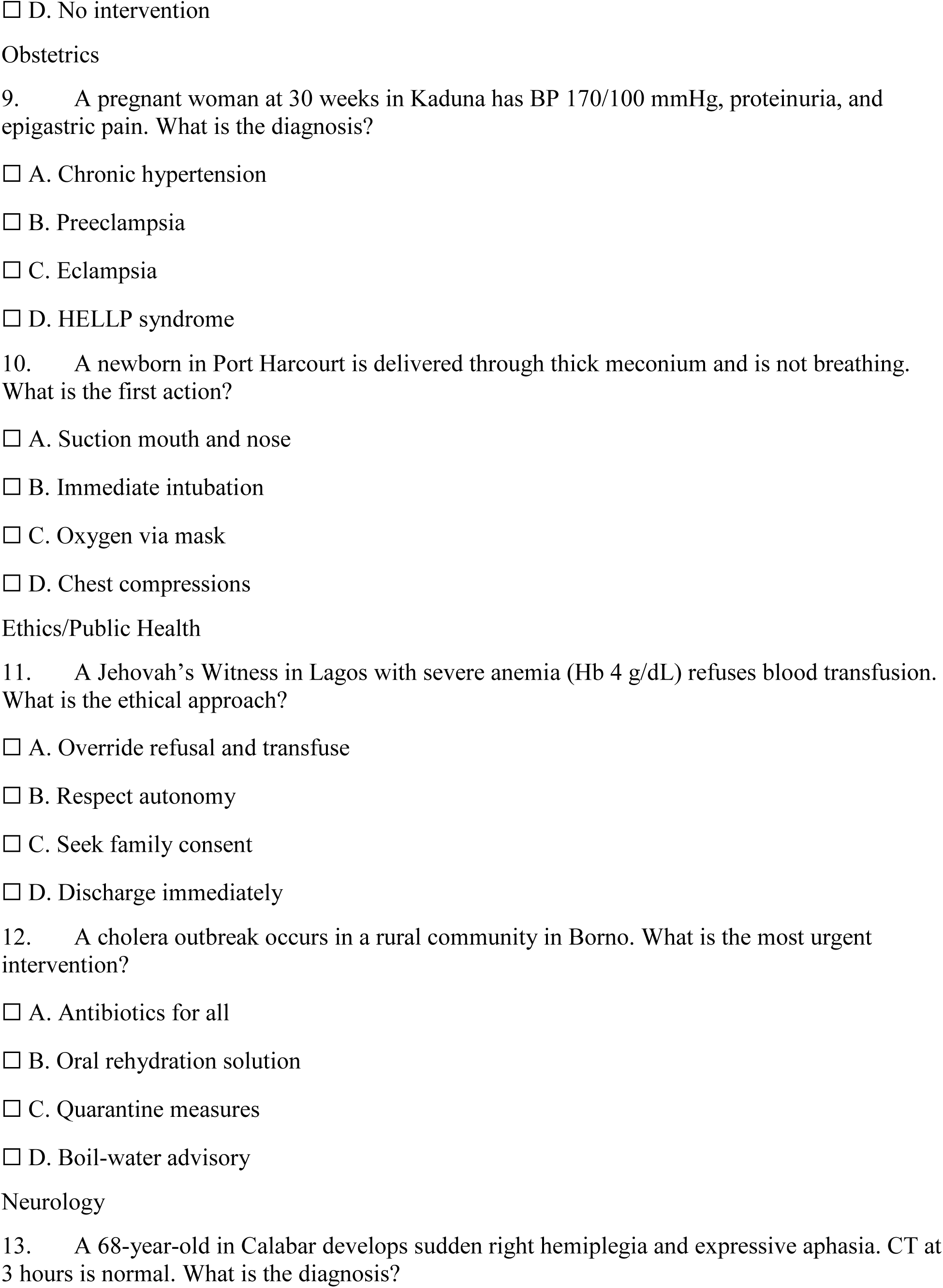
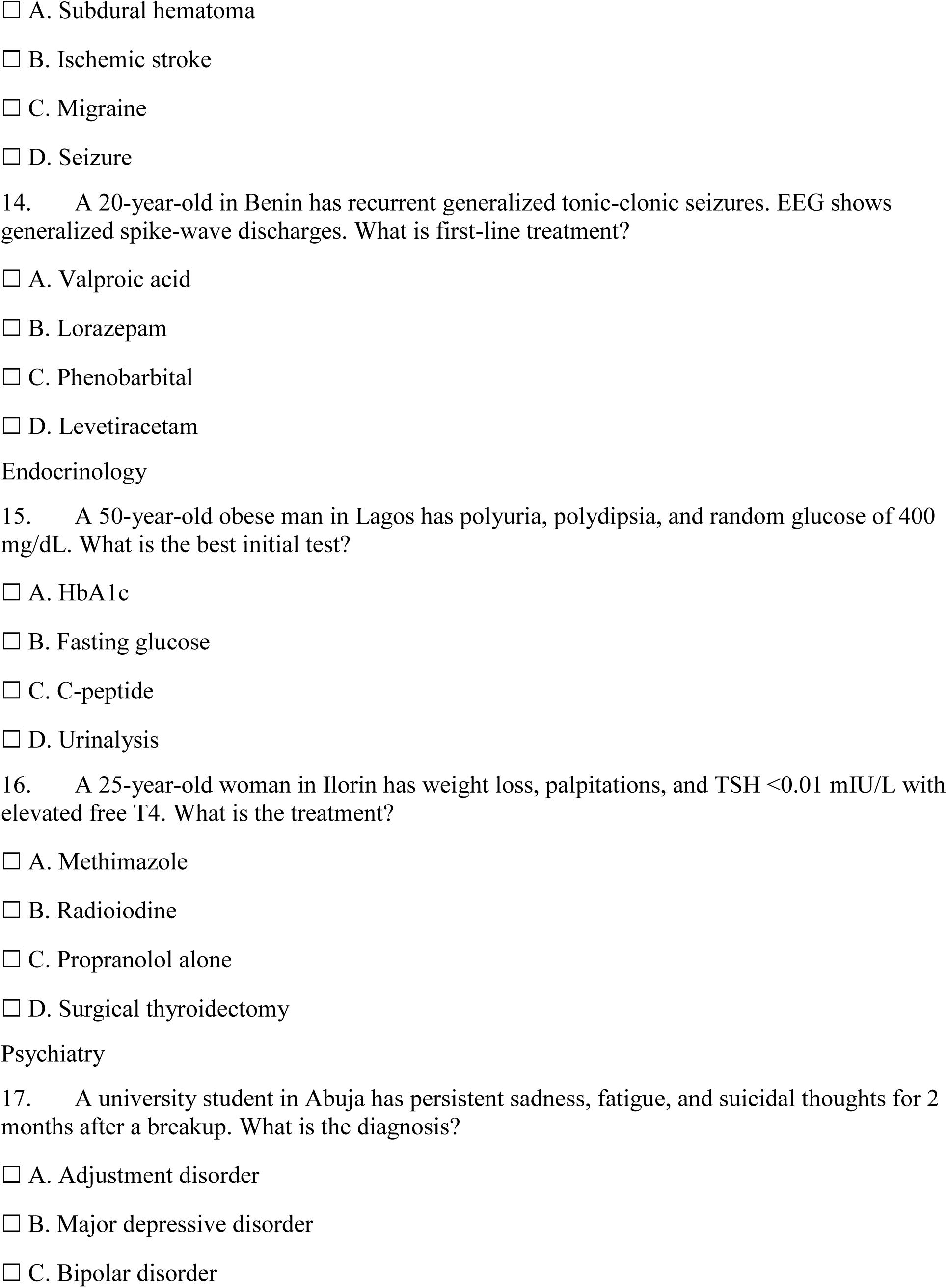
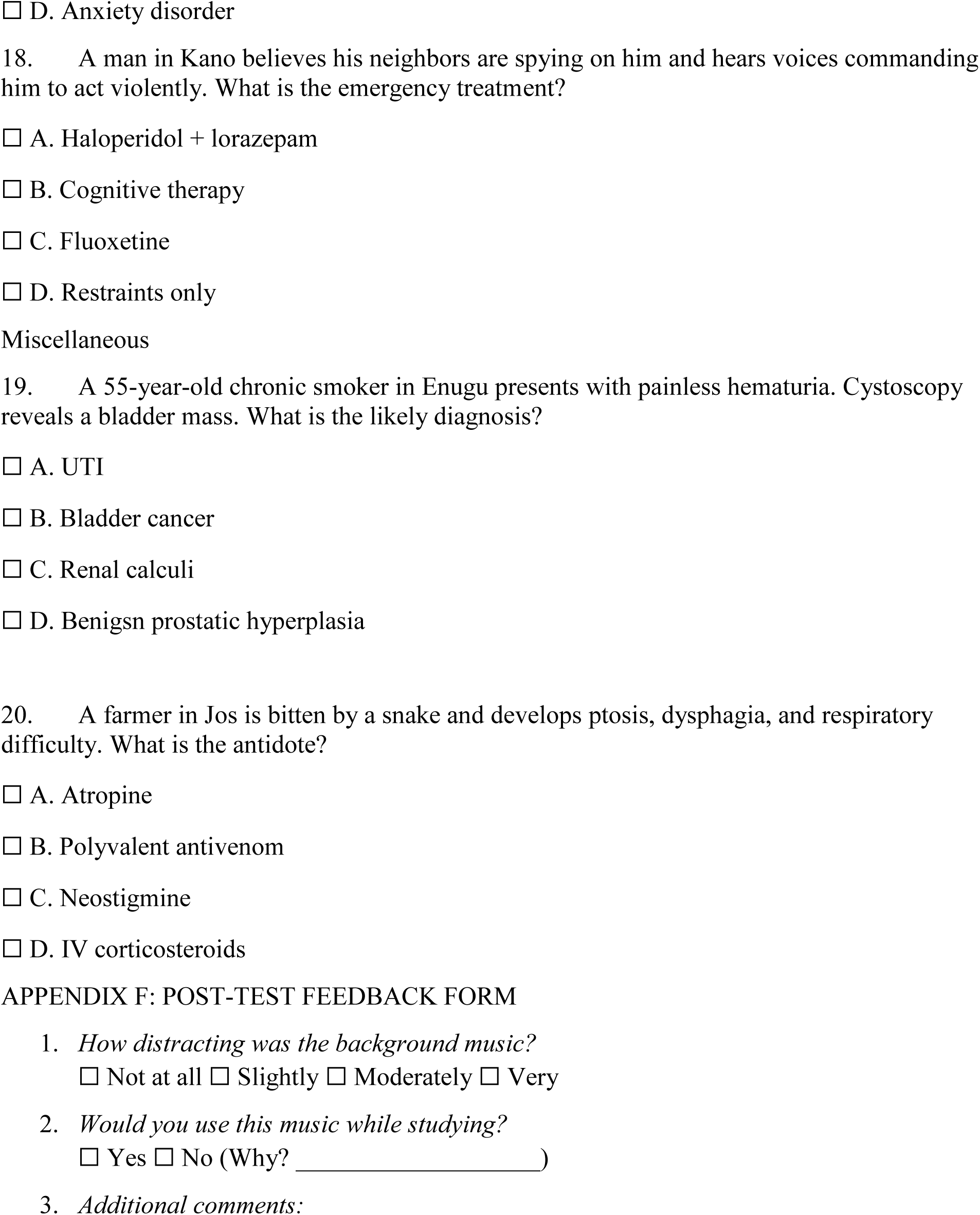

