## Supplemental Files for "IMPACT OF BACKGROUND IGBO HIGHLIFE MUSIC ON COGNITIVE PERFORMANCE AMONG CLINICAL MEDICAL STUDENTS: A COMPARATIVE STUDY OF MEMORY RECALL AND PROBLEM-SOLVING EFFICIENCY"

Principal Investigator: Chidi Anaenye
Institution: Nnamdi Azikiwe University, Awka
Contact: 07064298863

1. Study Purpose

This research examines how background music affects medical students' memory and problem-solving skills.

2. Procedures

- Duration: 3 sessions (45 mins each)
- Tasks: Memory tests, MCQs, Mood surveys
- Music Exposure: Random assignment to Highlife/Classical/Silence

3. Risks & Benefits

- Risks: Minimal (similar to studying with music)
- Benefits: Contributes to optimal study strategies

4. Confidentiality

- Data anonymized
- Secure storage

5. Voluntary Participation

✓ I consent voluntarily
✓ I may withdraw anytime
✓ I understand the purpose

Participant Signature: ____________________ Date: _________

Researcher Signature: ____________________ Date: _________

#### APPENDIX B: DEMOGRAPHIC SURVEY

DEMOGRAPHIC SURVEY

Study Title: *The Impact of Igbo Highlife Music on Cognitive Performance*

SECTION A: BASIC INFORMATION

1. Age: _____ years
2. Gender:
   ☐ Male
   ☐ Female
   ☐ Non-binary/Other (specify): _________
3. Ethnicity:
   ☐ Igbo
   ☐ Yoruba
   ☐ Hausa
   ☐ Other (specify): _________

SECTION B: ACADEMIC BACKGROUND

4. Medical School Year:
   ☐ 3rd Year
   ☐ 4th Year
   ☐ 5th Year
   ☐ 6th Year
5. Average Study Hours Per Day:
   ☐ <2 hours
   ☐ 2–4 hours
   ☐ >4 hours

SECTION C: MUSIC EXPOSURE & HABITS

6. Do you typically study with background music?
   ☐ Yes → *Genre most used:* _________
   ☐ No
7. How familiar are you with Igbo Highlife music?
   ☐ Not at all
   ☐ Slightly familiar
   ☐ Moderately familiar
   ☐ Very familiar
8. Formal Music Training (if any):
   ☐ None
   ☐ 1–5 years (Instrument: _________)
   ☐ >5 years (Instrument: _________)

SECTION D: HEALTH & LIFESTYLE

9. Do you have any diagnosed hearing impairments?
   ☐ Yes
   ☐ No
10. Sleep Quality (Past Week):
   ☐ Very poor
   ☐ Poor
   ☐ Fair
   ☐ Good
   ☐ Excellent

SECTION E: OPEN-ENDED RESPONSE

11. *What factors most distract you while studying?*

#### APPENDIX C: BASELINE MOOD ASSESSMENT (PANAS)

Instructions: Rate the extent you feel each emotion RIGHT NOW using this scale:

1 = Very Slightly or Not at All | 2 = A Little | 3 = Moderately | 4 = Quite a Bit | 5 = Extremely

| Item | 1 | 2 | 3 | 4 | 5 |
|---|---|---|---|---|---|
| 1. Interested | ○ | ○ | ○ | ○ | ○ |
| 2. Stressed | ○ | ○ | ○ | ○ | ○ |
| 3. Enthusiastic | ○ | ○ | ○ | ○ | ○ |
| 4. Anxious | ○ | ○ | ○ | ○ | ○ |
| 5. Focused | ○ | ○ | ○ | ○ | ○ |
| 6. Irritable | ○ | ○ | ○ | ○ | ○ |
| 7. Energetic | ○ | ○ | ○ | ○ | ○ |
| 8. Overwhelmed | ○ | ○ | ○ | ○ | ○ |
| 9. Alert | ○ | ○ | ○ | ○ | ○ |
| 10. Relaxed (reverse) | ○ | ○ | ○ | ○ | |

### APPENDIX D: MEMORY RECALL TEST

MEMORY RECALL TEST PROTOCOL

Title: *Advanced Medical Terminology Recall Assessment*
Time Allotted: 5 minutes (study) + 3 minutes (recall)
Instructions for Participants:

1. Study Phase (5 minutes):
   - Memorize the 20 terms and definitions displayed on the screen/printed sheet.
   - *No note-taking allowed*.
2. Distraction Task (2 minutes):
   - Perform a simple cognitive task (e.g., count backward from 100 by 7s).
3. Recall Phase (3 minutes):
   - Write down all terms you remember *verbatim* (synonyms not accepted).
   - *Optional*: For bonus points, add one key clinical detail per term (e.g., "Charcot Triad = cholangitis").

20 MEDICAL TERMS FOR MEMORY RECALL

1. Rhabdomyolysis
*Definition*: Skeletal muscle breakdown releasing myoglobin → acute kidney injury (CK >5,000 U/L).
*Key*: "Tea-colored urine," treat with aggressive IV hydration.

2. Charcot Triad
*Definition*: Fever + RUQ pain + jaundice = ascending cholangitis.
*Key*: Medical emergency requiring ERCP + antibiotics.

3. Osler Nodes
*Definition*: Tender subcutaneous nodules in infective endocarditis.
*Key*: Immune complex deposition (vs. Janeway lesions = non-tender).

4. Leukoerythroblastosis
*Definition*: Nucleated RBCs + immature WBCs on peripheral smear.
*Key*: Suggests bone marrow infiltration (e.g., metastatic cancer).

5. Hamman’s Sign
*Definition*: Crunching sound on auscultation (mediastinal emphysema).
*Key*: Pathognomonic for pneumomediastinum (e.g., post-esophageal rupture).

6. Pulsus Paradoxus

*Definition*: >10 mmHg drop in SBP during inspiration.
*Key*: Cardiac tamponade hallmark (also seen in severe asthma).

7. Grey-Turner's Sign
*Definition*: Flank ecchymosis in hemorrhagic pancreatitis.
*Key*: Retroperitoneal bleeding (Cullen's sign = periumbilical).

8. Obtundation
*Definition*: Reduced alertness with minimal response to stimuli.
*Key*: Between lethargy and stupor in consciousness spectrum.

9. Saddle PE
*Definition*: Pulmonary embolism straddling main pulmonary artery bifurcation.
*Key*: High mortality, may cause sudden cardiovascular collapse.

10. Angioedema
*Definition*: Bradykinin-mediated subcutaneous swelling (non-pitting).
*Key*: ACE inhibitor-induced cases don't respond to epinephrine.

11. Erythroderma
*Definition*: >90% body surface erythema with scaling.
*Key*: Life-threatening (e.g., Sézary syndrome, drug reactions).

12. Anion Gap Metabolic Acidosis
*Definition*: AG >12 due to unmeasured anions (e.g., lactate, ketones).
*Key*: MUDPILES mnemonic for causes.

13. Heberden's Nodes
*Definition*: Bony enlargements of DIP joints in osteoarthritis.
*Key*: Unlike Bouchard's nodes (PIP joints).

14. Torus Palatinus
*Definition*: Benign bony protrusion on hard palate.
*Key*: Asymptomatic; must distinguish from oral cancer.

15. Lymphadenopathy
*Definition*: Pathologic lymph node enlargement (>1 cm).
*Key*: "Shotty" nodes suggest viral etiology; rubbery = lymphoma.

16. Bronchiolitis Obliterans

*Definition*: Fibrotic small airway obstruction post-inflammation.
*Key*: "Popcorn lung" in popcorn factory workers (diacetyl exposure).
17. Erythema Migrans
*Definition*: Expanding targetoid rash of Lyme disease.
*Key*: Requires doxycycline even without serologic confirmation.

18. Chvostek's Sign
*Definition*: Facial twitching upon tapping facial nerve (hypocalcemia).
*Key*: Less specific than Trousseau's sign.

19. Asterixis
*Definition*: "Liver flap" from impaired ammonia metabolism.
*Key*: Negative myoclonus (brief loss of muscle tone).

20. Virchow's Triad
*Definition*: Endothelial injury + stasis + hypercoagulability = VTE.
*Key*: Underlies DVT/PE pathophysiology.

TEST ANSWER SHEET

SERIAL NO: ___________________ DATE: ___________
GROUP: [Igbo Highlife/Classical/Silence]

RECALL AS MANY TERMS AS POSSIBLE AND WRITE THEM DOWN IN THE FOLLOWING FORMAT (3 MINUTES):

1. ________________________________________ Key Detail: ___________________________

### APPENDIX E: PARALLEL MEMORY RECALL TEST

SERIAL NO: __________________ DATE: ___________

GROUP: [Igbo Highlife/Classical/Silence]

#### PARALLEL MEMORY RECALL TEST PROTOCOL

Title: *Advanced Medical Terminology Recall Assessment*
Time Allotted: 5 minutes (study) + 3 minutes (recall)

Instructions for Participants:

4. Study Phase (5 minutes):
   - Memorize the 20 terms and definitions displayed on the screen/printed sheet.
   - *No note-taking allowed*.
5. Distraction Task (2 minutes):
   - Perform a simple cognitive task (e.g., count backward from 100 by 7s).
6. Recall Phase (3 minutes):
   - Write down all terms you remember *verbatim* (synonyms not accepted).
   - *Optional*: For bonus points, add one key clinical detail per term (e.g., "Charcot Triad = cholangitis").

#### PARALLEL 20 MEDICAL TERMS FOR MEMORY RECALL

1. **Myoglobinuria**
   Definition: Presence of myoglobin in urine due to muscle breakdown.
   Key: Differentiate from hematuria (no RBCs on microscopy).

2. **Reynolds' Pentad**
   Definition: Charcot triad + hypotension + AMS (severe cholangitis).
   Key: Indicates septic shock requiring urgent intervention.

3. **Janeway Lesions**
   Definition: Non-tender macules on palms/soles in endocarditis.
   Key: Represent septic microemboli (vs. Osler nodes).

4. **Leukemoid Reaction**
   Definition: WBC >50,000/μL without malignancy.
   Key: Seen in severe infections, unlike true leukemia.

5. **Subcutaneous Emphysema**
   Definition: Air in subcutaneous tissues (crepitus on palpation).
   Key: Associated with pneumothorax or esophageal rupture.

6. **Pulsus Alternans**
   Definition: Alternating strong/weak pulses in severe LV dysfunction.
   Key: Poor prognostic sign in heart failure.

7. **Cullen's Sign**
   Definition: Periumbilical ecchymosis in hemorrhagic pancreatitis.
   Key: Indicates retroperitoneal hemorrhage.

8. **Stupor**
   Definition: State of near-unconsciousness, responsive only to pain.
   Key: More severe than obtundation.

9. **Massive PE**
   Definition: PE causing hypotension (SBP<90) or cardiac arrest.
   Key: Different from saddle PE (location vs. hemodynamics).

10. **Hereditary Angioedema**
    Definition: C1 esterase inhibitor deficiency causing swelling.
    Key: Bradykinin-mediated (unresponsive to antihistamines).

11. **Exfoliative Dermatitis**
    Definition: Generalized erythema and scaling (>90% BSA).
    Key: Life-threatening drug reaction or cutaneous lymphoma.

12. **Non-Anion Gap Metabolic Acidosis**
    Definition: Hyperchloremic acidosis with normal anion gap.
    Key: Causes include diarrhea, renal tubular acidosis.

13. **Bouchard's Nodes**
    Definition: Bony enlargements of PIP joints in OA.
    Key: Differentiate from rheumatoid nodules.

14. **Torus Mandibularis**
    Definition: Bony protrusion on mandibular lingual surface.
    Key: Benign anatomical variant.

15. **Lymphangitis**
    Definition: Inflammation of lymphatic vessels (red streaks).
    Key: Typically streptococcal infection.

16. **Bronchiectasis**

Definition: Irreversible bronchial dilation with chronic infection.

Key: "Tram-track" appearance on imaging.

17. **Erythema Nodosum**

Definition: Tender subcutaneous nodules on shins.

Key: Hypersensitivity reaction (sarcoidosis, TB, strep).

18. **Trousseau's Sign**

Definition: Carpal spasm after BP cuff inflation (hypocalcemia).

Key: More specific than Chvostek's sign.

19. **Myxedema Coma**

Definition: Life-threatening severe hypothyroidism.

Key: Treat with IV levothyroxine and glucocorticoids.

20. **Virchow's Node**

Definition: Left supraclavicular lymphadenopathy.

Key: May indicate gastric cancer metastasis.

#### APPENDIX F: PROBLEM-SOLVING MCQ (SAMPLE)

SERIAL NO: ___________________ DATE: ___________

GROUP: [Igbo Highlife/Classical/Silence]

Time Limit: 20 minutes (1 min/question)

Infectious Diseases

1. A 35-year-old man in Abuja presents with high fever, severe headache, and retro-orbital pain. Lab findings reveal leukopenia and thrombocytopenia (70,000/µL). What is the most likely diagnosis?
   ☐ A. Lassa fever
   ☐ B. Yellow fever
   ☐ C. Dengue
   ☐ D. Leptospirosis

2. A 7-year-old in Sokoto has fever, altered consciousness, and a petechial rash. CSF shows Gram-positive cocci in pairs. What is the immediate treatment?
   ☐ A. IV ceftriaxone
   ☐ B. Oral amoxicillin
   ☐ C. IV acyclovir
   ☐ D. IV quinine

Cardiology

3. A 50-year-old man with a history of smoking collapses at a gym in Enugu. ECG shows ventricular tachycardia without a pulse. What is the next step?
   ☐ A. IV lidocaine
   ☐ B. Defibrillation
   ☐ C. IV metoprolol
   ☐ D. CPR for 5 minutes

4. A 65-year-old diabetic man has crushing chest pain lasting 30 minutes. ECG shows ST elevation in leads II, III, and aVF. Which medication should be given first?
   ☐ A. Nitroglycerin
   ☐ B. Aspirin
   ☐ C. IV heparin
   ☐ D. Morphine

Pediatrics

5. A 3-month-old in Lagos has chronic diarrhea, foul-smelling stools, and poor weight gain. Sweat chloride is elevated. What is the diagnosis?
   ☐ A. Celiac disease
   ☐ B. Cystic fibrosis
   ☐ C. Lactose intolerance
   ☐ D. Giardiasis

6. A neonate in Kano has jaundice at 24 hours of life with bilirubin of 15 mg/dL. Mother is Rh-negative, baby is Rh-positive. What is the best initial management?
   ☐ A. Phototherapy
   ☐ B. Exchange transfusion
   ☐ C. IVIG
   ☐ D. Discontinue breastfeeding

Surgery/Trauma

7. A car accident victim in Ibadan has hypotension, abdominal distension, and a positive FAST scan. What is the next step?
   ☐ A. Abdominal ultrasound
   ☐ B. Exploratory laparotomy
   ☐ C. CT scan
   ☐ D. IV fluids only

8. A farmer in Owerri sustains a deep wound contaminated with soil. His last tetanus booster was 15 years ago. What is required?
   ☐ A. Tetanus toxoid only
   ☐ B. Both toxoid and immunoglobulin
   ☐ C. Antibiotics alone
   ☐ D. No intervention

Obstetrics

9. A pregnant woman at 30 weeks in Kaduna has BP 170/100 mmHg, proteinuria, and epigastric pain. What is the diagnosis?
   ☐ A. Chronic hypertension
   ☐ B. Preeclampsia
   ☐ C. Eclampsia
   ☐ D. HELLP syndrome

10. A newborn in Port Harcourt is delivered through thick meconium and is not breathing. What is the first action?
   ☐ A. Suction mouth and nose

☐ B. Immediate intubation
☐ C. Oxygen via mask
☐ D. Chest compressions

Ethics/Public Health

11. A Jehovah's Witness in Lagos with severe anemia (Hb 4 g/dL) refuses blood transfusion. What is the ethical approach?
☐ A. Override refusal and transfuse
☐ B. Respect autonomy
☐ C. Seek family consent
☐ D. Discharge immediately

12. A cholera outbreak occurs in a rural community in Borno. What is the most urgent intervention?
☐ A. Antibiotics for all
☐ B. Oral rehydration solution
☐ C. Quarantine measures
☐ D. Boil-water advisory

Neurology

13. A 68-year-old in Calabar develops sudden right hemiplegia and expressive aphasia. CT at 3 hours is normal. What is the diagnosis?
☐ A. Subdural hematoma
☐ B. Ischemic stroke
☐ C. Migraine
☐ D. Seizure

14. A 20-year-old in Benin has recurrent generalized tonic-clonic seizures. EEG shows generalized spike-wave discharges. What is first-line treatment?
☐ A. Valproic acid
☐ B. Lorazepam
☐ C. Phenobarbital
☐ D. Levetiracetam

Endocrinology

15. A 50-year-old obese man in Lagos has polyuria, polydipsia, and random glucose of 400 mg/dL. What is the best initial test?
☐ A. HbA1c
☐ B. Fasting glucose
☐ C. C-peptide
☐ D. Urinalysis

16. A 25-year-old woman in Ilorin has weight loss, palpitations, and TSH <0.01 mIU/L with elevated free T4. What is the treatment?
☐ A. Methimazole
☐ B. Radioiodine
☐ C. Propranolol alone
☐ D. Surgical thyroidectomy

Psychiatry

17. A university student in Abuja has persistent sadness, fatigue, and suicidal thoughts for 2 months after a breakup. What is the diagnosis?
☐ A. Adjustment disorder
☐ B. Major depressive disorder
☐ C. Bipolar disorder
☐ D. Anxiety disorder

18. A man in Kano believes his neighbors are spying on him and hears voices commanding him to act violently. What is the emergency treatment?
☐ A. Haloperidol + lorazepam
☐ B. Cognitive therapy
☐ C. Fluoxetine
☐ D. Restraints only

Miscellaneous

19. A 55-year-old chronic smoker in Enugu presents with painless hematuria. Cystoscopy reveals a bladder mass. What is the likely diagnosis?
☐ A. UTI
☐ B. Bladder cancer
☐ C. Renal calculi
☐ D. Benign prostatic hyperplasia

20. A farmer in Jos is bitten by a snake and develops ptosis, dysphagia, and respiratory difficulty. What is the antidote?
☐ A. Atropine
☐ B. Polyvalent antivenom
☐ C. Neostigmine
☐ D. IV corticosteroids

### APPENDIX G: PARALLEL TEST PROBLEM SOLVING ABILITY

SERIAL NO: ___________________ DATE: ___________

GROUP: [Igbo Highlife/Classical/Silence]

Parallel Test Problem-Solving Ability MCQ Bank (20 Items)

Time Limit: 20 minutes (1 min/question)

Infectious Diseases

1. A 35-year-old man in Abuja presents with high fever, severe headache, and retro-orbital pain. Lab findings reveal leukopenia and thrombocytopenia (70,000/μL). What is the most likely diagnosis?

☐ A. Lassa fever

☐ B. Yellow fever

☐ C. Dengue

☐ D. Leptospirosis

2. A 7-year-old in Sokoto has fever, altered consciousness, and a petechial rash. CSF shows Gram-positive cocci in pairs. What is the immediate treatment?

☐ A. IV ceftriaxone

☐ B. Oral amoxicillin

☐ C. IV acyclovir

☐ D. IV quinine

Cardiology

3. A 50-year-old man with a history of smoking collapses at a gym in Enugu. ECG shows ventricular tachycardia without a pulse. What is the next step?

☐ A. IV lidocaine

☐ B. Defibrillation

☐ C. IV metoprolol

☐ D. CPR for 5 minutes

4. A 65-year-old diabetic man has crushing chest pain lasting 30 minutes. ECG shows ST elevation in leads II, III, and aVF. Which medication should be given first?

☐ A. Nitroglycerin

☐ B. Aspirin

☐ C. IV heparin

☐ D. Morphine

Pediatrics

5. A 3-month-old in Lagos has chronic diarrhea, foul-smelling stools, and poor weight gain. Sweat chloride is elevated. What is the diagnosis?

☐ A. Celiac disease

☐ B. Cystic fibrosis

☐ C. Lactose intolerance

☐ D. Giardiasis

6. A neonate in Kano has jaundice at 24 hours of life with bilirubin of 15 mg/dL. Mother is Rh-negative, baby is Rh-positive. What is the best initial management?

☐ A. Phototherapy

☐ B. Exchange transfusion

☐ C. IVIG

☐ D. Discontinue breastfeeding

Surgery/Trauma

7. A car accident victim in Ibadan has hypotension, abdominal distension, and a positive FAST scan. What is the next step?

☐ A. Abdominal ultrasound

☐ B. Exploratory laparotomy

☐ C. CT scan

☐ D. IV fluids only

8. A farmer in Owerri sustains a deep wound contaminated with soil. His last tetanus booster was 15 years ago. What is required?

☐ A. Tetanus toxoid only

☐ B. Both toxoid and immunoglobulin

☐ C. Antibiotics alone

☐ D. No intervention

Obstetrics

9. A pregnant woman at 30 weeks in Kaduna has BP 170/100 mmHg, proteinuria, and epigastric pain. What is the diagnosis?

☐ A. Chronic hypertension

☐ B. Preeclampsia

☐ C. Eclampsia

☐ D. HELLP syndrome

10. A newborn in Port Harcourt is delivered through thick meconium and is not breathing. What is the first action?

☐ A. Suction mouth and nose

☐ B. Immediate intubation

☐ C. Oxygen via mask

☐ D. Chest compressions

Ethics/Public Health

11. A Jehovah's Witness in Lagos with severe anemia (Hb 4 g/dL) refuses blood transfusion. What is the ethical approach?

☐ A. Override refusal and transfuse

☐ B. Respect autonomy

☐ C. Seek family consent

☐ D. Discharge immediately

12. A cholera outbreak occurs in a rural community in Borno. What is the most urgent intervention?

☐ A. Antibiotics for all

☐ B. Oral rehydration solution

☐ C. Quarantine measures

☐ D. Boil-water advisory

Neurology

13. A 68-year-old in Calabar develops sudden right hemiplegia and expressive aphasia. CT at 3 hours is normal. What is the diagnosis?

☐ A. Subdural hematoma

☐ B. Ischemic stroke

☐ C. Migraine

☐ D. Seizure

14. A 20-year-old in Benin has recurrent generalized tonic-clonic seizures. EEG shows generalized spike-wave discharges. What is first-line treatment?

☐ A. Valproic acid

☐ B. Lorazepam

☐ C. Phenobarbital

☐ D. Levetiracetam

#### Endocrinology

15. A 50-year-old obese man in Lagos has polyuria, polydipsia, and random glucose of 400 mg/dL. What is the best initial test?

☐ A. HbA1c

☐ B. Fasting glucose

☐ C. C-peptide

☐ D. Urinalysis

16. A 25-year-old woman in Ilorin has weight loss, palpitations, and TSH <0.01 mIU/L with elevated free T4. What is the treatment?

☐ A. Methimazole

☐ B. Radioiodine

☐ C. Propranolol alone

☐ D. Surgical thyroidectomy

#### Psychiatry

17. A university student in Abuja has persistent sadness, fatigue, and suicidal thoughts for 2 months after a breakup. What is the diagnosis?

☐ A. Adjustment disorder

☐ B. Major depressive disorder

☐ C. Bipolar disorder

☐ D. Anxiety disorder

18. A man in Kano believes his neighbors are spying on him and hears voices commanding him to act violently. What is the emergency treatment?

☐ A. Haloperidol + lorazepam

☐ B. Cognitive therapy

☐ C. Fluoxetine

☐ D. Restraints only

Miscellaneous

19. A 55-year-old chronic smoker in Enugu presents with painless hematuria. Cystoscopy reveals a bladder mass. What is the likely diagnosis?

☐ A. UTI

☐ B. Bladder cancer

☐ C. Renal calculi

☐ D. Benigsn prostatic hyperplasia

20. A farmer in Jos is bitten by a snake and develops ptosis, dysphagia, and respiratory difficulty. What is the antidote?

☐ A. Atropine

☐ B. Polyvalent antivenom

☐ C. Neostigmine

☐ D. IV corticosteroids

APPENDIX F: POST-TEST FEEDBACK FORM

1. *How distracting was the background music?*
   ☐ Not at all ☐ Slightly ☐ Moderately ☐ Very
2. *Would you use this music while studying?*
   ☐ Yes ☐ No (Why? ____________________)
3. *Additional comments:*
